# Trans-ancestry genome-wide association study identifies novel genetic mechanisms in rheumatoid arthritis

**DOI:** 10.1101/2021.12.01.21267132

**Authors:** Kazuyoshi Ishigaki, Saori Sakaue, Chikashi Terao, Yang Luo, Kyuto Sonehara, Kensuke Yamaguchi, Tiffany Amariuta, Chun Lai Too, Vincent A Laufer, Ian C Scott, Sebastien Viatte, Meiko Takahashi, Koichiro Ohmura, Akira Murasawa, Motomu Hashimoto, Hiromu Ito, Mohammed Hammoudeh, Samar Al Emadi, Basel K Masri, Hussien Halabi, Humeria Badsha, Imad W Uthman, Xin Wu, Li Lin, Ting Lin, Darren Plant, Anne Barton, Gisela Orozco, Suzanne MM Verstappen, John Bowes, Alexander J MacGregor, Suguru Honda, Masaru Koido, Kohei Tomizuka, Yoichiro Kamatani, Hiroaki Tanaka, Eiichi Tanaka, Akari Suzuki, Yuichi Maeda, Kenichi Yamamoto, Satoru Miyawaki, Gang Xie, Jinyi Zhang, Chris Amos, Ed Keystone, Gertjan Wolbink, Irene van der Horst-Bruinsma, Jing Cui, Katherine P Liao, Robert J Carroll, Hye-Soon Lee, So-Young Bang, Katherine A Siminovitch, Niek de Vries, Lars Alfredsson, Solbritt Rantapää-Dahlqvist, Elizabeth W Karlson, Sang-Cheol Bae, Robert P Kimberly, Jeffrey C Edberg, Xavier Mariette, Tom Huizinga, Philippe Dieudé, Matthias Schneider, Martin Kerick, Joshua C Denny, The Biobank Japan Project, Koichi Matsuda, Keitaro Matsuo, Tsuneyo Mimori, Fumihiko Matsuda, Keishi Fujio, Yoshiya Tanaka, Atsushi Kumanogoh, Matthew Traylor, Cathryn M Lewis, Stephen Eyre, Huji Xu, Richa Saxena, Thurayya Arayssi, Yuta Kochi, Katsunori Ikari, Masayoshi Harigai, Peter K Gregersen, Kazuhiko Yamamoto, S. Louis Bridges, Leonid Padyukov, Javier Martin, Lars Klareskog, Yukinori Okada, Soumya Raychaudhuri

**Author notes:** **Corresponding authors:** Yukinori Okada and Soumya Raychaudhuri. these authors contributed equally (Kazuyoshi Ishigaki, Saori Sakaue, Chikashi Terao).

## Abstract

Trans-ancestry genetic research promises to improve power to detect genetic signals, fine-mapping resolution, and performances of polygenic risk score (PRS). We here present a large-scale genome-wide association study (GWAS) of rheumatoid arthritis (RA) which includes 276,020 samples of five ancestral groups. We conducted a trans-ancestry meta-analysis and identified 124 loci (*P* < 5 × 10^-8^), of which 34 were novel. Candidate genes at the novel loci suggested essential roles of the immune system (e.g., *TNIP2* and *TNFRSF11A*) and joint tissues (e.g., *WISP1*) in RA etiology. Trans-ancestry fine mapping identified putatively causal variants with biological insights (e.g., *LEF1*). Moreover, PRS based on trans-ancestry GWAS outperformed PRS based on single-ancestry GWAS and had comparable performance between European and East Asian populations. Our study provides multiple insights into the etiology of RA and improves genetic predictability of RA.

## Main text

Rheumatoid arthritis (RA) is an autoimmune disease in which the immune system attacks the synovium in the joints, leading to chronic tissue inflammation, joint destruction, and disability. While recent therapeutic developments now alter the course of disease, RA mechanisms have yet to be fully elucidated and a cure has yet to be identified. RA can be divided into two major subtypes (seropositive and seronegative RA) based on the presence or absence of RA-specific serum antibodies (rheumatoid factor or anti-citrullinated peptide antibodies)^1^. Since RA is highly heritable^2–4^, genetic research has the potential to advance our understanding of its pathology. Indeed, previous studies successfully identified candidates of causal alleles, genes, pathways, and cell types^2,5–7^. For example, previous studies suggested that CD4^+^ effector T cells play a central role and the T cell receptor signaling pathway drives autoimmunity in RA^7–10^.

Trans-ancestry genetic research has multiple advantages over single-ancestry analysis. First, genome-wide association study (GWAS) in a single ancestry can be underpowered to detect signals from a causal allele with low allele frequency in that ancestry. As notable examples, the causal alleles can be ancestry-specific, as shown in studies for other complex diseases^11–13^. Having multiple ancestries with different allele frequency spectrum can improve the power. Second, single-ancestry GWAS are hampered by the specific linkage disequilibrium (LD) structure in that ancestry, which could obscure the ability to effectively fine-map an associated locus^14,15^. Trans-ancestry GWAS can improve fine-mapping resolution by leveraging the distinct LD structures in each ancestry^15–17^. Third, PRS generally has limited transferability across ancestries. For example, when the PRS model is developed based on GWAS in European (EUR) populations, PRS performs poorly in non-EUR populations^18^. PRS based on trans-ancestry GWAS can potentially improve its performance in multiple ancestries^19,20^; this is a clinically important topic since PRS can benefit patients via precision medicine. Although many RA genetic studies were conducted in non-EUR populations^2,3,17,21–24^, they were relatively small in the sample sizes, and much larger research efforts have focused on EUR populations^6,25–32^.

Here, we report a large-scale trans-ancestry GWAS of RA, including individuals of EUR, East Asian (EAS), African (AFR), South Asian (SAS), and Arabian (ARB) ancestries. While seropositive and seronegative RA are associated with phenotypic differences, they have shared heritability^33^, and their risk alleles appear to be similar outside of the major histocompatibility complex (MHC) locus^34^. Therefore, we initially focused on all RA, and then we restricted cases to seropositive patients. After identifying novel loci, we conducted fine-mapping to elucidate potential molecular mechanism of risk alleles. We examined the extent to which genetic signals are shared across ancestries while also investigating ancestry-specific genetic signals. We developed PRS models using our GWAS results and compared their performances across all ancestries. Our study provides multiple insights into the etiology of RA and highlights the importance and further needs of diversifying the ancestral background of GWAS participants.

## RESULTS

### Trans- and single-ancestry GWAS

We included 37 cohorts comprising 35,871 RA patients and 240,149 control individuals of EUR, EAS, AFR, SAS, and ARB ancestry (**Figure 1a**; **Supplementary Table 1** and **2**); 22,350 cases and 74,823 controls in 25 EUR cohorts; 11,025 cases and 162,608 controls in eight EAS cohorts; 999 cases and 1,108 controls in two AFR cohorts; 986 cases and 1,258 controls in one SAS cohort; and 511 cases and 352 controls in one ARB cohort. RA-specific serum antibodies were measured in 31,963 (89%) of cases; among them 27,448 (86%) were seropositive and 4,515 (14%) were seronegative (**Supplementary Table 1; Methods**). To confirm the diversity of ancestral backgrounds, we projected each individual’s genotype into principal component (PC) space which was calculated using all individuals in 1000 Genomes Project Phase 3 (1KG Phase3). We further conducted uniform manifold approximation and projection (UMAP) using their top 20 PC scores. This revealed finer scale ancestral structure, and confirmed that our study represented many 1KG Phase 3 ancestries (**Figure 1b** and **1c**; **Extended Data Figure 1**).

**Figure 1.**
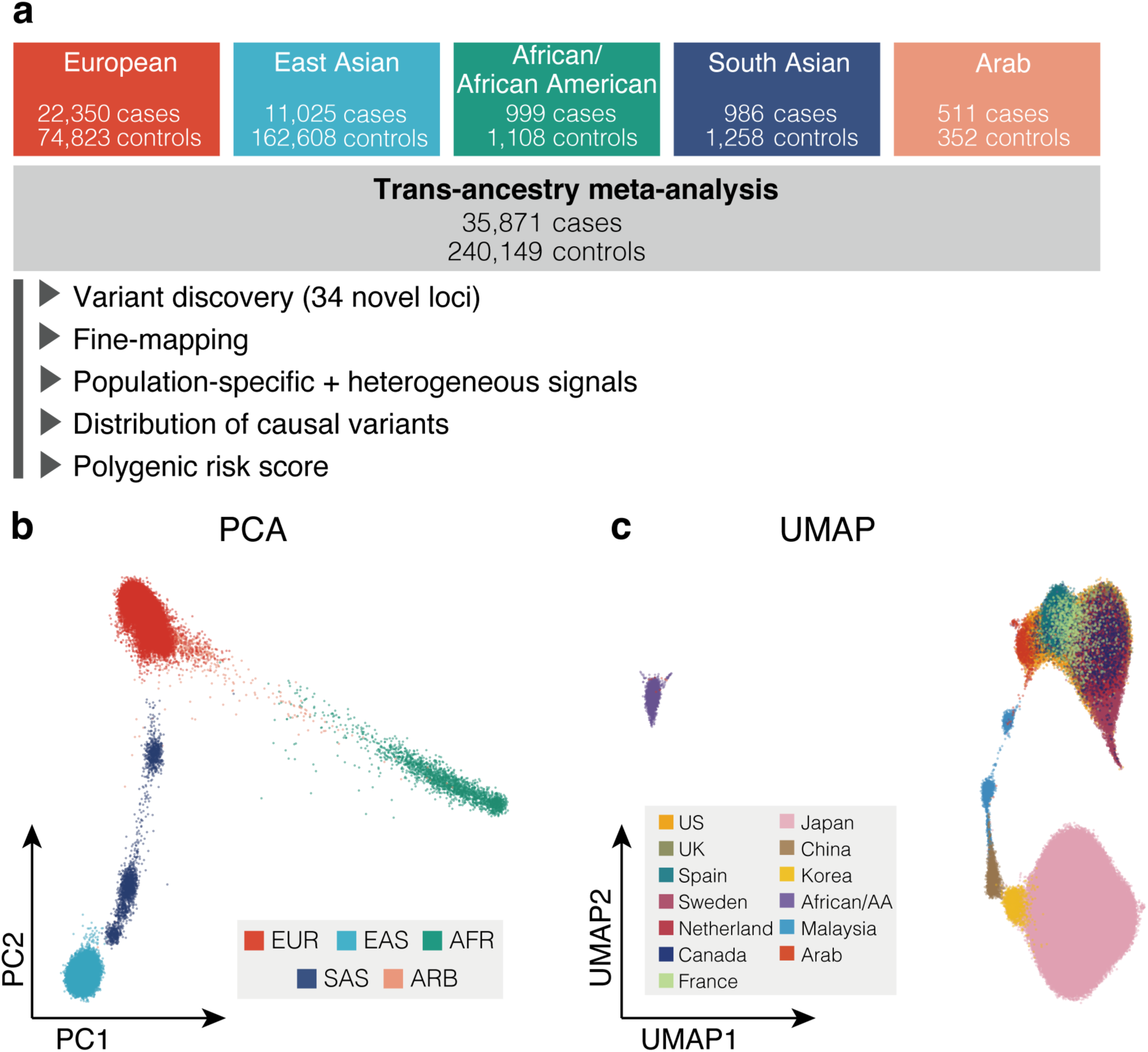
Diverse ancestral background in this GWAS participants. **(a)** Study design of this GWAS. The total number of cases and controls are provided. **(b)** PCA plot of all GWAS samples. We projected each individual’s imputed genotype into a PC space which was calculated using all individuals in 1KG Phase3. The samples are colored by its ancestry group. **(c)** UMAP plots of all GWAS samples. We conducted UMAP analysis using their top 20 PC scores. The samples in a cohort were colored by the country/region-level group of that cohort (**Supplementary Table 1**). When a cohort recruited participants from multiple countries, we did not plot its samples.

After quality control and imputation, we conducted GWAS in each cohort by logistic regression (**Methods**). We calculated genomic inflation using all variants outside of the MHC locus and observed little evidence of statistical inflation (mean of lambda = 1.01; S.D = 0.04; **Supplementary Table 1**). Primary analysis included all cases, while we restricted cases to seropositive patients in a secondary analysis.

We then conducted a meta-analysis using all cohorts across five ancestries by the inverse-variance weighted fixed effect model (trans-ancestry GWAS). We observed almost identical effect sizes between this trans-ancestry GWAS and the previously reported 100 RA risk alleles^2^ (Pearson’s *r* = 0.98 and *P* = 2.8 × 10^-82^; **Supplementary Figure 1**; **Supplementary Table 3**). We detected 108 genome-wide significant loci (*P* < 5 × 10^−8^) in this trans-ancestry study: 106 autosomal loci and two loci on the X chromosome (**Supplementary Table 4** and **5**). For ancestries with multiple cohorts (EUR, EAS, and AFR), we also conducted a meta-analysis within each ancestry by the same strategy (EUR-, EAS-, and AFR-GWAS). EUR-GWAS identified three additional autosomal loci which were not significant in trans-ancestry GWAS (**Supplementary Table 4**). GWAS of seropositive RA additionally detected 13 autosomal loci (**Supplementary Table 4**). In total, we detected significant signals at 122 autosomal loci outside of the MHC locus and two loci on the X chromosome (**Supplementary Table 4** and **5**; we provided Manhattan and QQ plots in **Supplementary Figure 2**). Among these 122, 34 autosomal loci were novel (**Table 1**). Notably, 25 novel loci had not been implicated in any other autoimmune diseases (**Supplementary Table 4; Methods**).

**Table 1.**
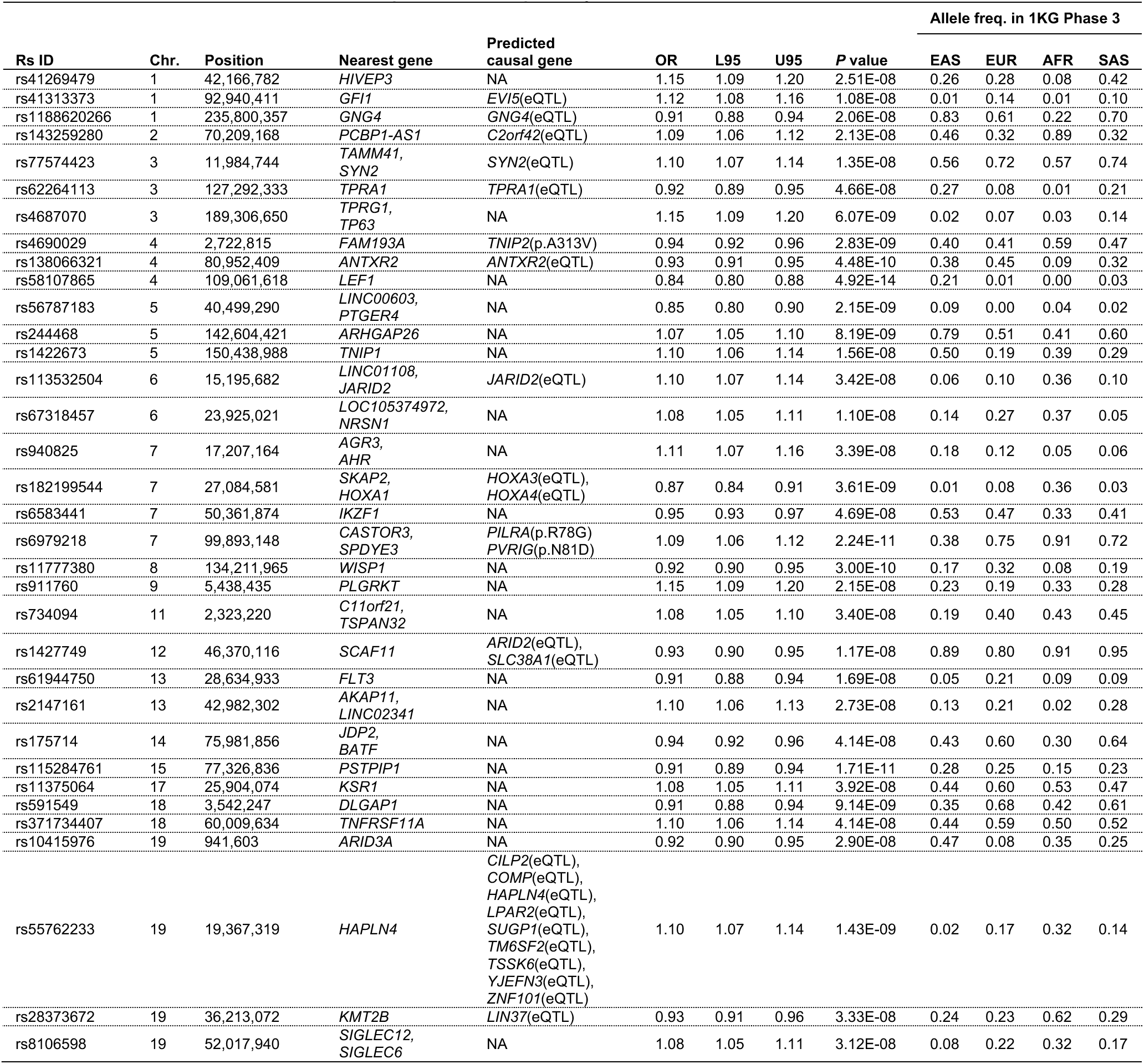
Novel RA risk loci detected in this study. Statistics in the GWAS setting with the lowest *P* values were provided (see **Supplementary Table 4** for details). The genomic coordinate is according to GRCh37. Predicted causal gene, predicted molecular consequences using eQTL or non-synonymous variants (see **Supplementary Table 8-10** for details); OR, odds ratio (the effect allele is the alternative allele); L95, lower 95% confidence interval; U95, upper 95% confidence interval; allele freq., allele frequency of the effect allele.

To quantify the heritability, we analyzed our GWAS results using stratified-linkage disequilibrium score regression (S-LDSC)^10^ (**Supplementary Table 2**). Since S-LDSC assumes GWAS has samples from a single ancestral background and a sufficient sample size, we restricted this analysis to EUR- and EAS-GWAS. The heritability explained by non-MHC common variants was similar between EUR and EAS; the liability scale heritability was 0.14 (S.E. = 0.01) for EUR and 0.13 (S.E. = 0.01) for EAS (**Methods**). LDSC also confirmed that the amount of potential bias in the GWAS results was minimal; LDSC’s intercept = 1.03 for EUR and 1.02 for EAS (**Supplementary Table 2**).

### Fine-mapping analysis

We fine-mapped these 122 autosomal loci using approximate Bayesian factor (ABF)^35^ (**Methods**). The 95% credible sets included only one variant at seven loci (**Figure 2a**). Of these seven, six have not been reported in prior studies that conducted trans-ancestry fine-mapping of RA^17,36^ (**Supplementary Table 6**). The 95% credible sets included less than ten variants at 43 loci (**Figure 2a**). We identified 35 fine-mapped variants with posterior inclusion probability (*PIP*) greater than 0.5, which agree with and largely subsume prior fine-mapping results^6,17,36^; in addition, nine novel loci are represented (**Figure 2b; Supplementary Figure 3; Supplementary Table 6**). The proportion of non-synonymous variants was higher in the credible set variants with high *PIP* (*PIP* > 0.5) than low *PIP* (odds ratio (OR) = 9.3; one-sided Fisher exact test *P* = 0.02; **Figure 2c**).

**Figure 2.**
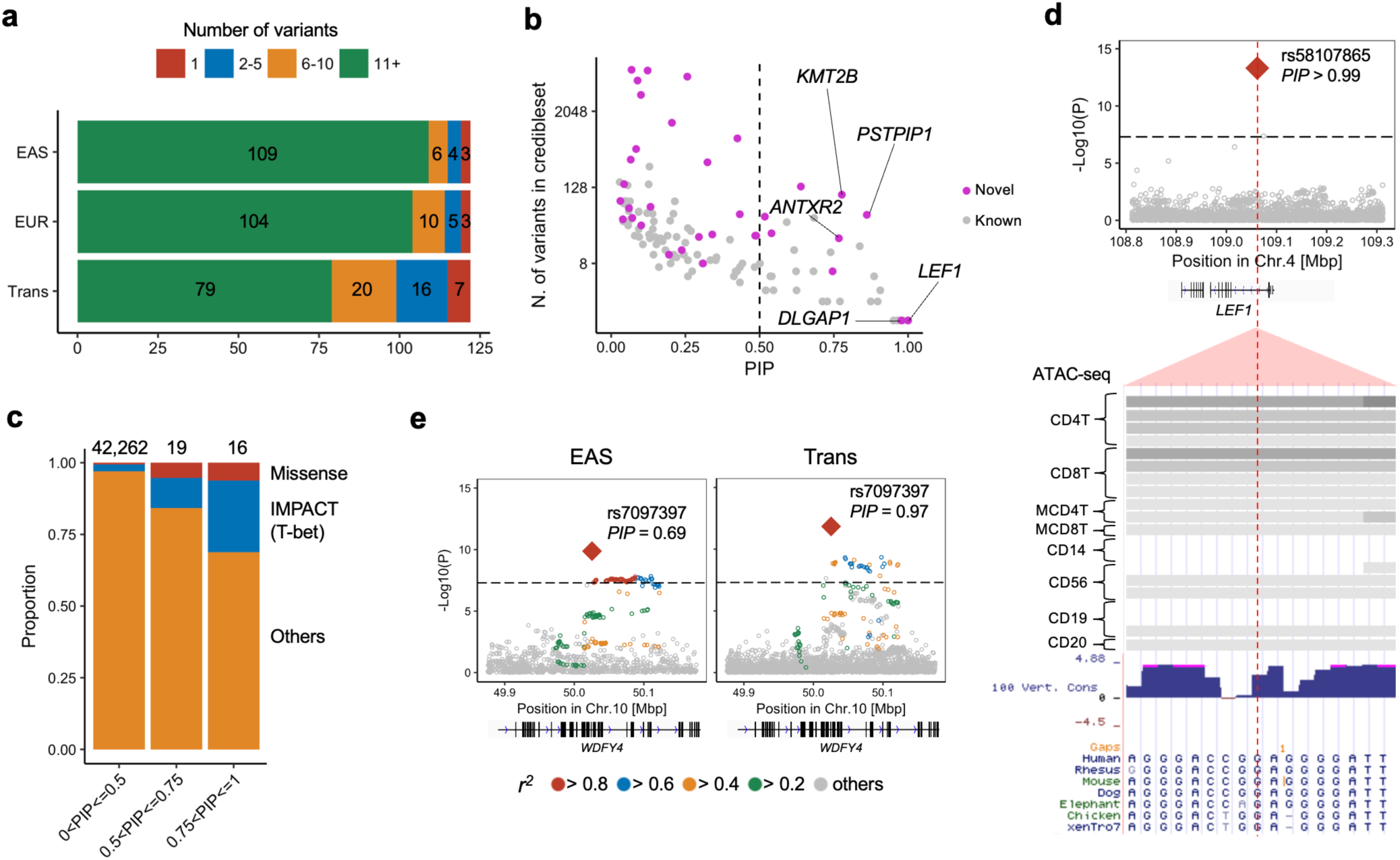
Fine-mapping analysis identified candidates of causal variants. **(a)** Among 122 autosomal loci analyzed, we counted the number of loci whose 95% credible set size was in a specified range. The results from EAS-, EUR- and trans-ancestry GWAS are provided. **(b)** The *PIP* of the lead variant and the size of 95% credible set at the 122 autosomal loci analyzed. The name of novel loci whose *PIP* was greater than 0.75 are labeled. We used trans-ancestry GWAS results. **(c)** In each range of *PIP* (the total number of variants were provided on the top), we calculated the proportion of non-synonymous variants or variants with high IMPACT score (CD4^+^ T cell T-bet annotation > 0.5). **(d)** A fine-mapped variant at the *LEF1* locus within CD4^+^ T cell specific open chromatin regions. *P* values of trans-ancestry GWAS are provided with dense view of immune cell ATAC-seq data (density indicate the read coverage) and vertebrate conservation data from UCSC genome browser (http://genome.ucsc.edu). **(e)** *P* values in the *WDFY4* locus in EAS- and trans-ancestry GWAS. We provide *r^2^* between each variant and the lead variant (rs7097397) by different colors. For trans-ancestry GWAS, we used intersection of LD variants in EUR and EAS ancestries.

We quantified the 95% credible set variants within open chromatin regions in 18 hematopoietic populations using gchromVAR software^37^. Consistent with previous analyses, we observed the strongest enrichment in CD4^+^ T cells (*P* = 5.4 × 10^-4^; **Extended Data Figure 2**). For example, rs58107865 at the *LEF1* locus (*PIP* > 0.99), rs7731626 at the *ANKRD55* locus (*PIP* > 0.99), and rs10556591 at the *ETS1* locus (*PIP* = 0.84) are located within CD4^+^ T cell-specific open chromatin regions (*Z* score > 2; **Supplementary Table 6**; **Methods**). Among them, rs58107865 is a novel risk variant and suggested the importance of regulatory T cells (T-reg) in RA biology (**Figure 2d**); LEF1 synergizes with FOXP3 to reinforce the gene networks essential for T-regs^38^. These results recapitulated a critical role of CD4^+^ T cells, especially T-regs, in RA biology.

As expected, compared with single-ancestry GWAS, trans-ancestry GWAS produced smaller sized credible sets and higher *PIP* (one-sided paired Wilcoxon text *P* < 3.1 × 10^-11^ and < 1.1 × 10^-9^, respectively) (**Figure 2a**; **Supplementary Figure 3**). For example, the *WDFY4* locus included 6,391 variants in the EUR 95% credible set, 64 variants in the EAS set, but only one in the trans-ancestry set, a missense variant of *WDFY4* (rs7097397; R1816Q; **Figure 2e**). Using a down-sampling experiment, we confirmed that this benefit was due to diversified LD structures as well as the increased sample size (**Supplementary Figure 3; Supplementary Note**).

### Conditional analysis

We conducted conditional analyses in each cohort to explore associations independent from the lead variants and meta-analyzed the results using the same strategy. We detected 24 independent signals at 21 loci (*P* < 5.0 × 10^−8^; **Supplementary Table 7**). Consistent with previous results^6,31^, we observed the largest number of independent associations at the *IL2RA*, *TYK2*, and *TNFAIP3* loci, where we observed three independent alleles (**Extended Data Figure 3**). The first and second lead variants at the *TYK2* locus were missense variants predicted to have damaging effects on *TYK2* protein function (SIFT score < 0.01; **Supplementary Table 8**).

In the *IL6R* locus, the conditional analysis identified two variants, rs12126142 (the first lead variant) and rs4341355 (the second lead variant), that were weakly correlated with each other but independently associated with RA (*r*^2^ = 0.23 and 0.15 in EUR and EAS of 1KG Phase 3, respectively; **Supplementary Table 7**). Interestingly, their protective alleles (rs12126142-A and rs4341355-C) almost always create a haplotype with the risk allele of the other variant (**Extended Data Figure 4**). Hence, the conditional analysis disentangled the independent yet mutually attenuating signals (**Figure 3a**). By analyzing expression quantitative trait loci (eQTL) and splicing quantitative trait loci (sQTL) in three immune cells from Blueprint consortium (CD4^+^ T cells, monocytes, and neutrophils)^39^, we found that rs12126142 and rs4341355 are likely to affect *IL6R* transcripts via different mechanisms. GWAS signals conditioned on rs4341355 colocalized with sQTL signals in monocytes (posterior probability of colocalization estimated by coloc software^40^ (*PP_coloc_*) > 0.99; **Figure 3b; Supplementary Table 9**); this sQTL signal corresponds to a previously-reported splicing isoform of soluble IL6R^41^. On the other hand, GWAS signals conditioned on rs12126142 colocalized with eQTL signals in CD4^+^ T cells (*PP_coloc_* = 0.97; **Figure 3b; Supplementary Table 9**). Therefore, our results suggested that both splicing and total expression of *IL6R* independently influence the RA genetic risk.

**Figure 3.**
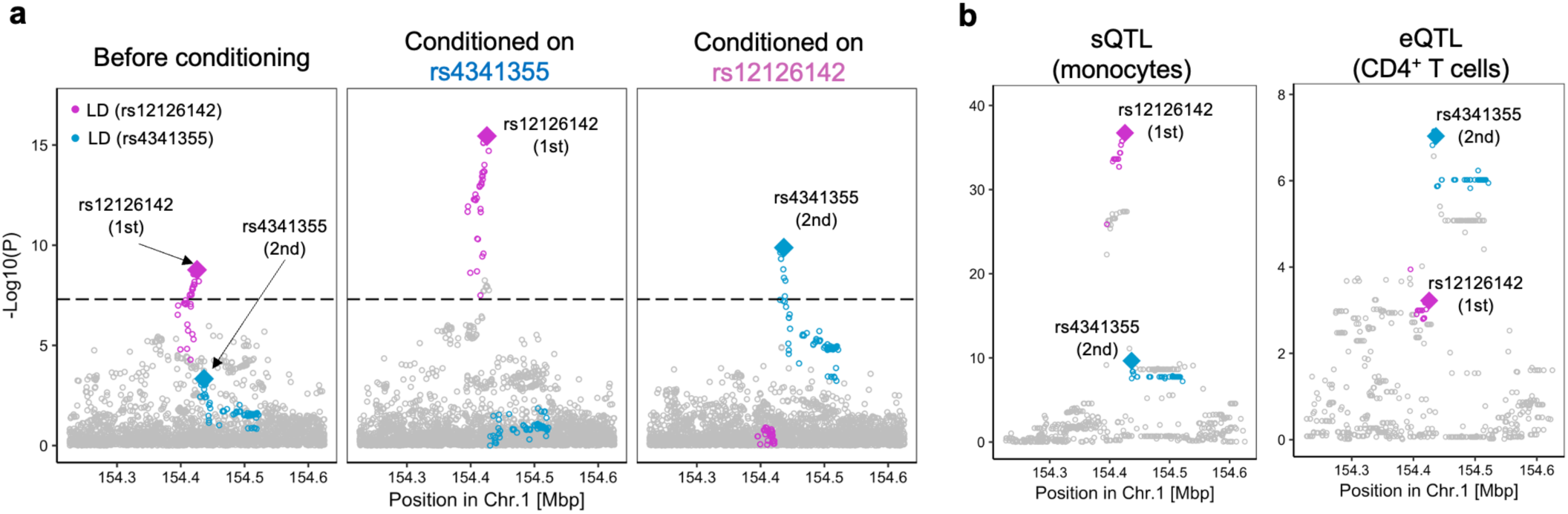
Splicing and total expression of *IL6R* jointly contribute to RA risk. **(a)** The 1^st^ lead variant (rs12126142; red) and the 2^nd^ lead variant (rs4341355; blue) are mutually attenuating their signals (controlling the effect of the other increased their signals). Conditional analysis was conducted in each cohort and the results were meta-analyzed using the inverse-variance weighted fixed effect model. We used trans-ancestry GWAS results. **(b)** *P* values of sQTL signals of *IL6R* (phenotype id: ENSG00000160712.8.17_154422457 in Blueprint dataset) and eQTL signals of *IL6R* (total expression of *IL6R*). We highlighted variants in LD with the lead variant by red or blue (*r^2^* > 0.6 in both EUR and EAS ancestries).

Our conditional analyses also suggested interesting biology at the *PADI4-PADI2* locus. We found two independent associations at this locus: esv3585367 (the first lead variant at a *PADI4* intron) and rs2076616 (the second lead variant at a *PADI2* intron), consistent with previous studies^17,42^ (**Figure 4a; Supplementary Table 7**). In sQTL results from the Blueprint consortium^39^, both *PADI4* and *PADI2* sQTL signals in neutrophils were colocalized with corresponding GWAS associations (*PP_coloc_* = 0.98 and 0.79, respectively; **Supplementary Table 9**), suggesting that alternative splicing of *PADI4* and *PADI2* likely increases the RA risk.

**Figure 4.**
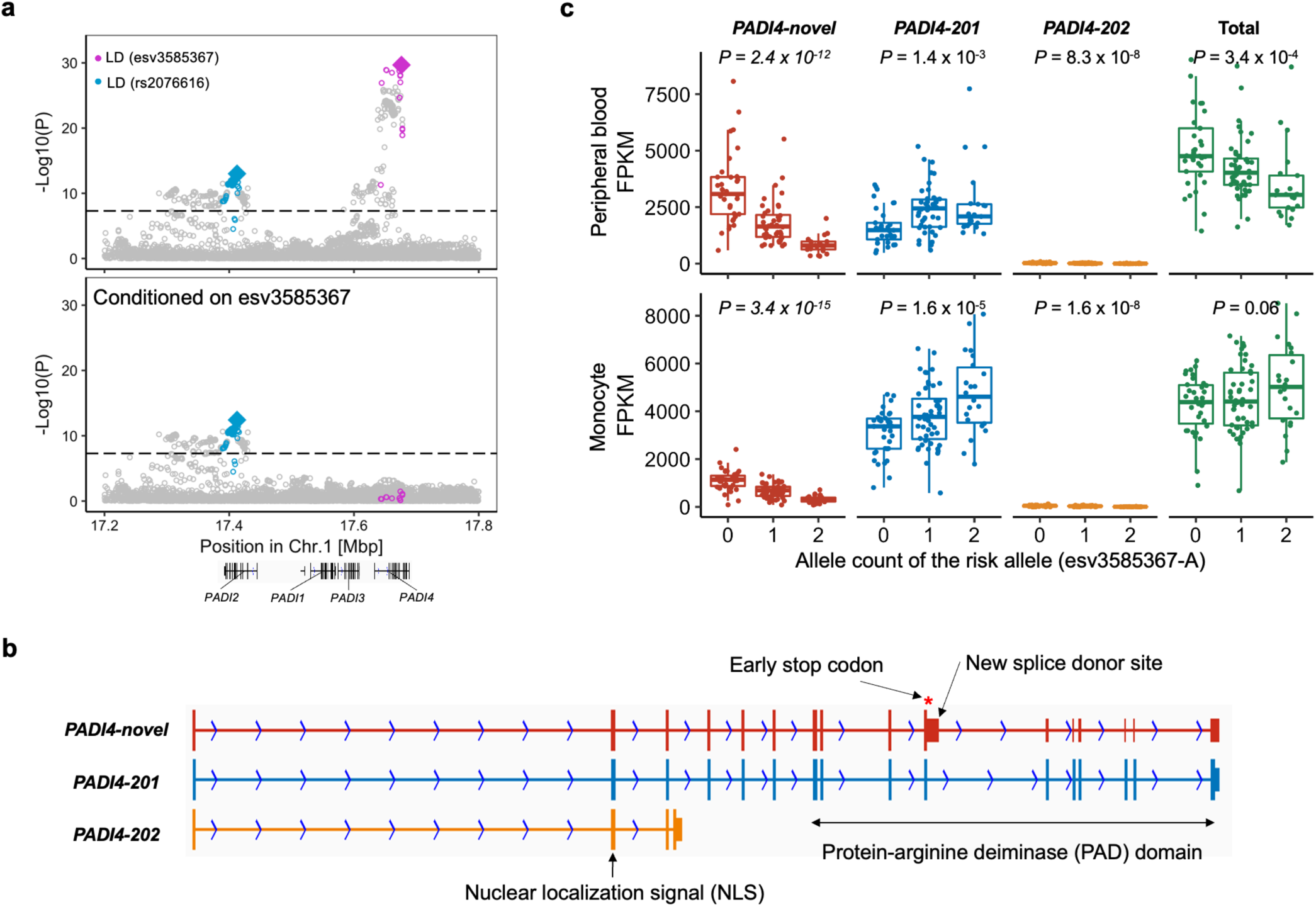
Splicing of *PADI4* contributes to RA risk. **(a)** Conditional analyses identified two independent associations at the *PADI4* locus: esv3585367 (red) and rs2076616 (blue). We used trans-ancestry GWAS results. We highlighted variants in LD with the lead variant by red or blue (*r*^2^ > 0.6 in both EUR and EAS ancestries). **(b)** A novel *PADI4* splice isoform confirmed by long-read sequencing datasets. *PADI4*-novel, a novel isoform we identified. *PADI4*-201, a functional isoform. *PADI4-novel* has an elongation of exon 10 which leads to an early stop codon and a truncated PAD domain at the C-terminus. PAD domain is an essential catalytic domain^46^, and highly conserved across other *PADI* genes. **(c)** The total expression and the expression of three isoforms of *PADI4* were plotted with the imputed dosages of the risk allele (esv3585367-A). The isoform structures were shown in **(b)**. We analyzed a RNA-seq dataset of 105 Japanese healthy individuals reported in a previous study^5^. We used peripheral blood leukocytes (neutrophils are its main component) and monocytes, both have high *PADI4* expression levels. *P* values from linear regression are provided. Within each boxplot, the horizontal lines reflect the median, the top and bottom of each box reflect the interquartile range (IQR), and the whiskers reflect the maximum and minimum values within each grouping no further than 1.5 × IQR from the hinge.

*PADI4* is critical for RA because it encodes an enzyme that citrullinates proteins, the main target for autoantibody in RA^23,43–45^. However, unlike *IL6R* with two functionally distinct isoforms^41^, full picture of *PADI4* splicing isoforms has not been extensively studied. To elucidate detailed molecular biology at the *PADI4* locus, we generated long-read sequencing datasets and inspected full-length *PADI4* transcripts. We identified a novel and probably non-functional splice isoform that produces a truncated protein-arginine deiminase (PAD) domain, an essential catalytic domain with two calcium-binding sites^46^ (**Figure 4b**). Next, we differentially quantified *PADI4* isoforms using RNA-seq data from 105 Japanese donors^5^. We found that the risk allele (esv3585367-A) was associated with the decrease of the non-functional novel isoform and the increase of the functionally intact isoform (**Figure 4c**). Notably, the allelic effect on the total expression, which had been conventionally used for eQTL studies, was not predictive of that on the functional isoform (the right panel in **Figure 4c**). Together, our analysis provided a novel genetic mechanism of *PADI4* and highlighted the importance of thoroughly investigating splice isoforms at the risk loci using long-read sequencing.

### Candidate causal genes at the associated loci

We next inferred the possible molecular consequences of all 148 detected variants: 124 lead variants (including two variants on the X chromosome) and 24 secondary variants detected by the conditional analysis.

We first focused on coding variants in LD with the lead variants in this GWAS (*r^2^* > 0.6 both in EUR and EAS samples in 1KG Phase 3; **Methods**). We found missense variants that may drive genetic signals at two novel loci (**Table 1**; **Supplementary Table 8**). An example is rs2269495 (A313V of *TNIP2*), in LD with a lead variant rs4690029 (*r^2^* = 0.65 and 0.89 in EUR and EAS of 1KG Phase 3, respectively). rs2269495 is predicted to have a damaging effect on TNIP2 protein function (SIFT score = 0.02; **Supplementary Table 8**). The protein product of *TNIP2* interacts with A20 (encoded by *TNFAIP3*) and inhibits NF-κB activation induced by TNF. Mice with a defective mutant *TNIP2* displayed intestinal inflammation and hypersensitivity to experimental colitis^47^. In addition, *TNIP1*, a homolog of *TNIP2,* was identified as one of the novel loci in this GWAS (**Supplementary Table 4**). Together, these results suggested that *TNIP1* and *TNIP2* are novel candidate causal genes of RA. Combined with the well-established *TNFAIP3* locus (ref^48^; **Supplementary Table 4**), these findings further supported the importance of the TNFAIP3-axis in RA biology.

We next inferred the possible molecular consequences using QTLs. We analyzed eQTL and sQTL in three immune cells from Blueprint consortium (CD4^+^ T cells, monocytes, and neutrophils) and multiple tissues from GTEx consortium^39,49^. We found colocalizing QTL signals at 11 novel loci (*PP_coloc_* > 0.7; **Table 1; Supplementary Table 9** and **10**). Several novel loci with colocalizing QTL signals suggested the biology of the non-immune systems, including joint tissues. For example, the risk allele of rs55762233 was associated with the increased expression of *CILP2* (*PP_coloc_* = 0.82), which encodes a component of the cartilage extracellular matrix. Its homologous gene, *CILP1*, was recently reported as one of the candidate autoantigens of RA^50^. Therefore, the protein product of *CILP2* might also have a critical role in RA biology.

We then searched for other biologically plausible genes which might explain novel signals and found several genes whose importance was supported by previous knowledge (**Table 1; Supplementary Table 4**). First, *TNFRSF11A* encodes RANK, a key regulator of osteoclast. Its ligand *RANKL* has been investigated as a potential therapeutic target^51,52^. *TNFRSF11A* is a causal gene for several bone-related Mendelian disorders^53–55^. Second, *WISP1* encodes a protein essential for osteoblast differentiation and bone formation^56,57^. In addition, *WISP1* is highly expressed in *HLA-DRA^hi^* inflammatory sublining fibroblasts, which are dramatically expanded and pathogenic in RA synovium^58^. Third, *FLT3* encodes a tyrosine kinase that regulates hematopoiesis and knocking out of whose ligand suppressed arthritis in model mice^59^. A damaging variant of *FLT3* was suggested to be associated with RA (*P* = 4.3 × 10^−4^) and other autoimmune diseases^60^.

### Differences and similarities of genetic risk across ancestries

We next searched for ancestry-specific signals at 122 autosomal loci. We defined ancestry-specificity when the lead variant in each locus is monomorphic in EUR or EAS samples of 1KG Phase 3. We found five EUR-specific signals: rs2476601 (a *PTPN22* missense variant), rs9826420 located in *STAG1* intronic region, rs7943728 (a *FADS2* eQTL), and rs34536443 and rs12720356 (both *TYK2* missense variants). EAS-GWAS also identified an EAS-specific signal at the *TYK2* locus: rs55882956, another *TYK2* missense variant. We thus detected two EUR-specific and one EAS-specific signal at the *TYK2* locus (**Extended Data Figure 5**). All these ancestry-specific signals were also reported by previous studies^2,25,61^. This study was underpowered to detect specific signals in non-EUR and non-EAS ancestries (**Supplementary Figure 4** and **Supplementary Note**). Although ancestry-specific signals are relatively few, they include predominantly large effect size variants, many of which are missense, and hence they are valuable resources to understand the etiology of RA.

Although we found these ancestry-specific signals, this study showed that genetic signals are generally shared across ancestries. We compared effect sizes between EUR-GWAS with non-EUR-GWASs at the 30 fine-mapped variants (*PIP* > 0.5; **Methods**). We found that the effect sizes were strongly correlated among five ancestries (Pearson’s *r* = 0.56-0.91; **Supplementary Figure 5**; **Supplementary Note**). In addition, we targeted genome-wide variants and tested the trans-ancestry genetic correlation between EUR- and EAS-GWAS using popcorn software^62^ (we restricted this analysis to EUR- and EAS-GWAS to avoid a biased correlation estimate caused by the small sample size). We again found a strong correlation (0.64 (S.E.=0.08); *P* = 4.4 × 10^-17^; *P* value reported is for a test that the correlation is different from 0).

### Genetic risk differences between seropositive and seronegative RA

The presence or absence of autoantibodies (rheumatoid factor and anti-citrullinated peptide antibodies) defines two major subgroups of RA: seropositive and seronegative RA^1^. We tested the differences in genetic signals between them at the 122 significant autosomal loci. Although their effect sizes were significantly correlated in general (Pearson’s *r* = 0.76; *P* = 3.2 × 10^-23^), we found significant heterogeneity in effect size estimates at the nine loci: *CCR6, CTLA4, NFKBIE-TMEM151B, PADI4, PTPN22, RAD51B, SMIM20-RBPJ, TNFRSF14-AS1*, and *UBASH3A* (*P_het_* < 0.05/122; **Extended Data Figure 6**). Effect size estimates were larger in seropositive RA at all the nine loci, and these findings suggested etiological differences between seropositive and seronegative RA. For example, *CCR6* has critical roles in B cell antibody production^63,64^. Together, these findings suggested generally shared genetic risks between seropositive and seronegative RA outside of the MHC locus, although substantial differences exist around biologically relevant genes.

### Genome-wide distributions of heritability

To acquire insights into RA biology, we estimated the heritability enrichments within gene regulatory regions using S-LDSC^10^, a method to infer the genome-wide distribution of all causal variants irrespective of their effect sizes. We again restricted this analysis to EUR- and EAS-GWAS. We utilized 707 IMPACT regulatory annotations, which reflect comprehensive cell-type-specific transcription factor (TF) activities^65^. Briefly, IMPACT probabilistically annotates each nucleotide genome-wide on a scale from 0 to 1, and we considered genomic regions scoring in the top 5% of each annotation. We detected significant enrichments in 114 annotations in either of EUR and EAS (*P* < 0.05/707 = 7.1 × 10^-5^; **Figure 5a** and **Supplementary Table 11**). The amount of heritability explained by each annotation was highly concordant between EUR and EAS (Pearson’s *r* = 0.92; *P* = 1.3 × 10^-290^; **Figure 5b**), and we did not observe significant heterogeneities between EUR and EAS estimates (*P_het_* > 0.05/707 = 7.1 × 10^-5^). Among annotations with significant enrichments, the one which explained the largest fraction of EUR heritability was CD4^+^ T cell T-bet annotation (90%; S.E. = 11%). This annotation explained 94% (S.E. = 12%) of EAS heritability consistent with analyses on previous GWAS results^66^. Although four out of 114 significant annotations were derived from non-immune cells, controlling the effect of CD4^+^ T cell T-bet annotation canceled out all four enrichments. We also analyzed 396 histone mark annotations, but they were less enriched in RA heritability than CD4^+^ T cell T-bet annotation (**Extended Data Figure 7; Supplementary Table 12; Supplementary Note**). In addition to identifying candidate critical TFs in RA pathology, these results also suggested that the distribution of causal variants is shared between EUR and EAS.

**Figure 5.**
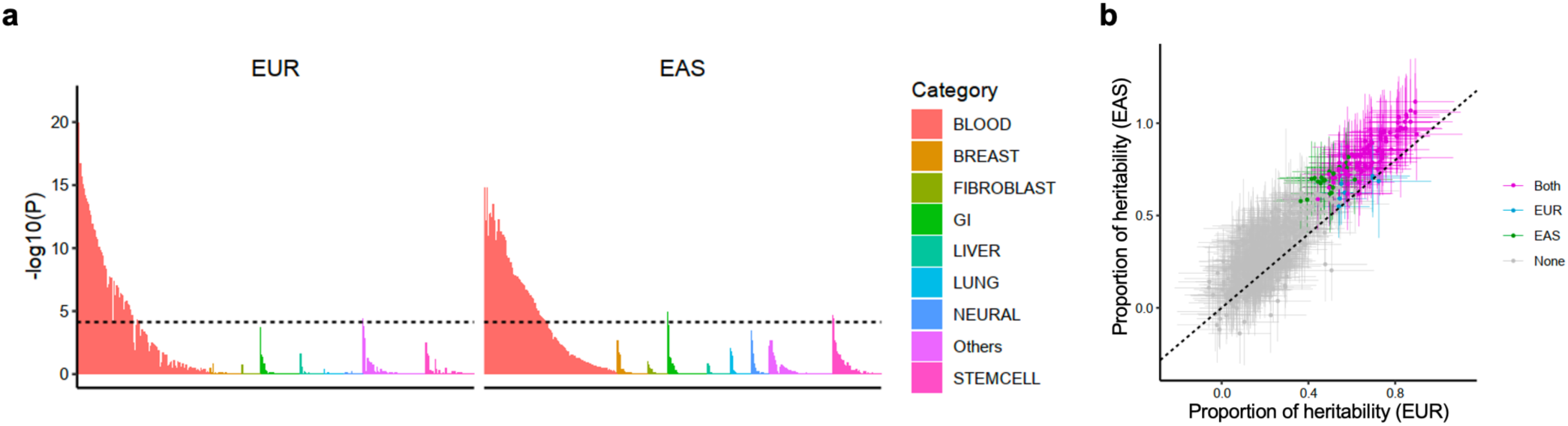
S-LDSC analysis suggested similar causal variant distributions in EUR- and EAS-GWAS. **(a)** EUR- and EAS-GWAS results were analyzed by S-LDSC using 707 IMPACT annotations. *P* value indicates significance of non-zero tau (per variant heritability) of each annotation. Each annotation was colored by its cell type category. Horizontal dashed line indicates Bonferroni-corrected *P* value threshold (0.05/707 = 7.1 × 10^-5^). **(b)** The estimate and its 95% confidence interval of the heritability proportion explained by top 5% of IMPACT annotations. When a heritability enrichment was significant (*P* < 0.05/707 = 7.1 × 10^-5^), that annotation was colored by the type of GWAS (“Both” indicates the annotation was significant both in EUR- and EAS-GWAS).

We next tested whether the findings in S-LDSC analysis can be recapitulated in fine-mapped variants from genome-wide significant loci. We analyzed credible set variants and found that high *PIP* variants (> 0.5) were enriched in high IMPACT score variants for CD4^+^ T cell T-bet annotation (> 0.5), compared with low *PIP* variants (**Figure 2c**; **Extended Data Figure 8**; OR = 8.7; one-sided Fisher exact test *P* = 1.5 × 10^-4^). We found six variants which possess high *PIP* and high IMPACT score, and one of them was a novel association at the intronic region of *LEF1* (rs58107865; **Supplementary Table 6**). Together, both polygenic and fine-mapped signals support the critical roles of CD4^+^ T cell’s T-bet activity in RA pathology.

### PRS performance across five ancestries

Our results showed that trans-ancestry GWAS can detect causal variants more efficiently than single-ancestry GWAS and these causal variants are strongly enriched within the CD4^+^ T cell T-bet annotation. Therefore, we hypothesized that trans-ancestry GWAS and CD4^+^ T cell T-bet annotation can improve PRS performances in non-EUR populations. To test this, we developed PRS models with six different conditions using combinations of two components: i) two variant selection settings (we used all variants or variants within the top 5% of the CD4^+^ T cell T-bet annotation and refer PRS based on each of them as standard or functionally-informed PRS, respectively) and ii) three GWAS settings (we used trans-ancestry, EUR-, or EAS-GWAS and refer PRS based on each of them as trans-ancestry, EUR-, or EAS-PRS, respectively). We designed our study so that there were no overlapping samples; when we evaluated the PRS performance in a given cohort, we re-conducted meta-analysis excluding that cohort to develop PRS models (**Figure 6a**; **Methods**). We defined the performance of PRS by phenotypic variance (pseudo-*R^2^*) explained by PRS. We tested PRS performances using ten different *P* value thresholds. We then selected the threshold with the best performance in each of the six PRS conditions separately and utilized this threshold for the following analyses.

**Figure 6.**
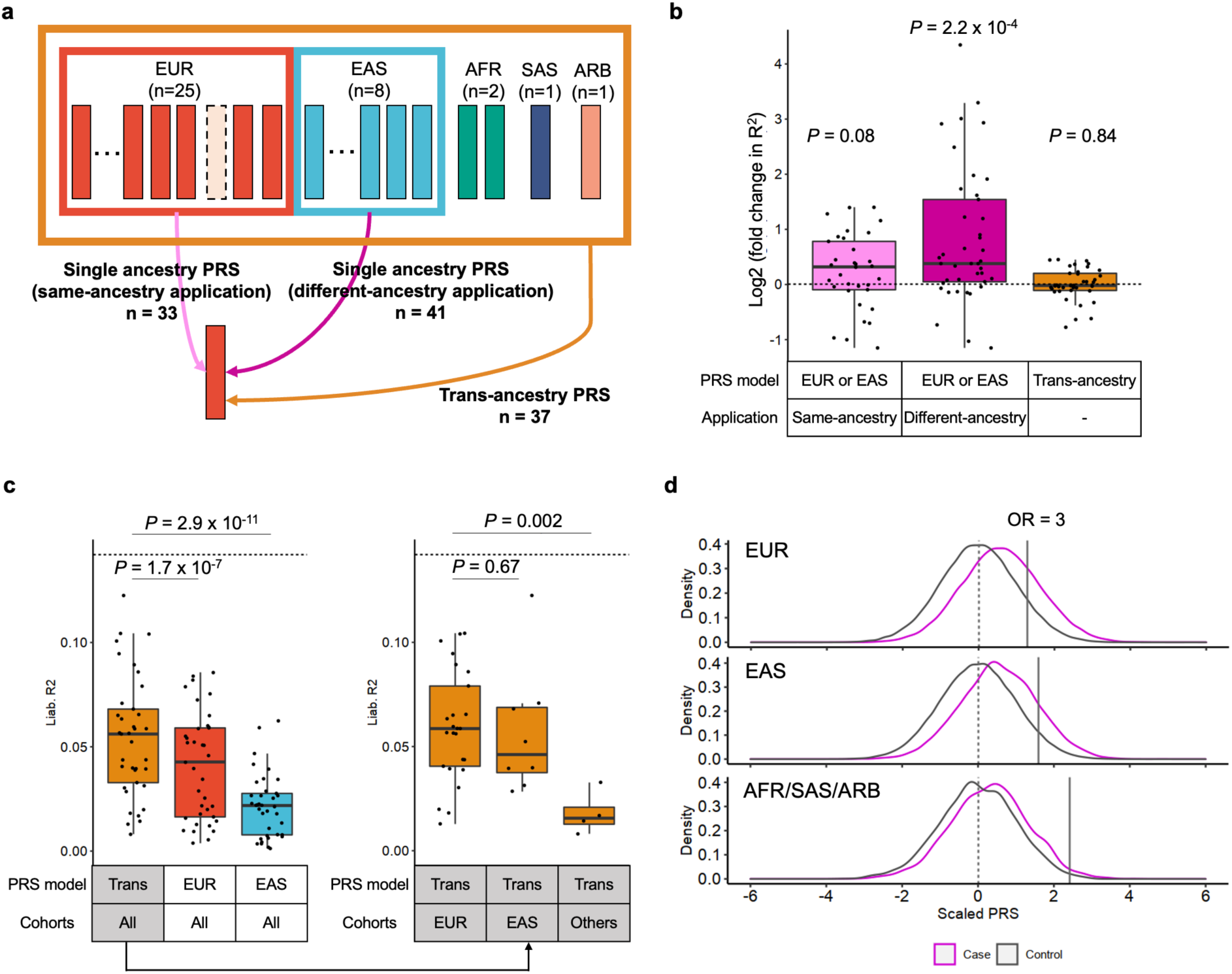
Functional annotation and trans-ancestry GWAS improved PRS performances. **(a)** The strategy of our PRS analysis. We utilized PRS based on three GWAS settings; two single-ancestry PRS (EUR- and EAS-PRS) and a trans-ancestry PRS. Single ancestry PRS had two types of applications; same-ancestry application of PRS (EUR-PRS applied to EUR cohorts or EAS-PRS applied to EAS cohorts) and different-ancestry application of PRS (EUR-PRS applied to non-EUR cohorts or EAS-PRS applied to non-EAS cohorts). When we applied a PRS model to a cohort included in a GWAS setting, we re-conducted the meta-analysis excluding that cohort to avoid overlapped samples. The numbers of each application are provided. **(b)** The improvements in PRS performance (*pseudo-R^2^*) by CD4^+^ T cell T-bet IMPACT annotation. Fold change indicates *R^2^* in functionally-informed PRS divided by *R^2^* in the standard PRS. We compared three conditions: same-ancestry application of single ancestry PRS (n=33), different-ancestry application of single ancestry PRS (n=41), and trans-ancestry PRS (n=37). One-sided sign test *P* value is provided. **(c)** The performance (liability-scale R^2^) of three different PRS models. The results of three PRS models in all cohorts are shown on the left panel. The results of trans-ancestry PRS in three cohort groups are shown on the right panel. The differences in *R^2^* were assessed by Wilcoxon test. In all conditions, CD4^+^ T cell T-bet IMPACT annotation was utilized to select variants. **(d)** PRS distribution differences between case and controls. Trans-ancestry PRS with CD4^+^ T cell T-bet IMPACT annotation was used. In each cohort, PRS was scaled using mean and SD of the control samples, and individual level data were merged across cohorts in an ancestral group. For a given PRS value at the right tail of PRS distribution, we compared the case-control ratios between individuals whose PRS is higher than that value and individuals whose PRS is lower than that value, and calculated the odds ratio (OR). The PRS values with OR = 3 are shown by solid vertical lines. Within each boxplot, the horizontal lines reflect the median, the top and bottom of each box reflect the interquartile range (IQR), and the whiskers reflect the maximum and minimum values within each grouping no further than 1.5 × IQR from the hinge.

We first evaluated the variant selection settings used for PRS (standard and functionally-informed PRS). Consistent with our recent study^65^, functionally-informed PRS improved the application of PRS constructed from different ancestries (EUR-PRS applied to non-EUR cohorts or EAS-PRS applied to non-EAS cohorts). Functionally-informed PRS increased *R^2^* by 2.7 fold on average, and we observed the improvement in 32 of 41 applications (one-sided sign test *P* = 2.2 × 10^-4^; **Figure 6b**; **Supplementary Table 13**). On the other hand, also consistent with our recent study^65^, this improvement was relatively small in the application of PRS constructed from the same ancestry (EUR-PRS applied to EUR cohorts or EAS-PRS applied to EAS cohorts). Functionally-informed PRS increased *R^2^* by 1.3 fold on average, and we observed the improvement in 21 out of 33 applications (one-sided sign test *P* = 0.08; **Figure 6b; Supplementary Table 13**). PRS based on single-ancestry GWAS is affected by LD structures of GWAS participants’ ancestral backgrounds; this can reduce performance in prediction when we apply this PRS to ancestries with different LD structures. These results confirmed that functionally-informed PRS can mitigate this problem. Expectedly, for trans-ancestry PRS, which prioritizes causal variants over variants solely associated through linkage, the benefit of functionally-informed PRS was very subtle; functionally-informed PRS increased *R^2^* only by 1.02 fold on average, and we observed the improvement only in 16 out of 37 applications (one-sided sign test *P* = 0.84; **Figure 6b**; **Supplementary Table 13**). Since CD4^+^ T cell T-bet annotation always had beneficial or neutral effects on PRS performances, we used functionally-informed PRS for the following analyses.

We next evaluated the GWAS settings used for PRS (trans-ancestry, EUR-, and EAS-PRS). Consistent with a recent study^14^, trans-ancestry PRS outperformed EUR-PRS and EAS-PRS; mean *R^2^* across 37 cohorts were 0.054, 0.041, and 0.022, in trans-ancestry, EUR-, and EAS-PRS, respectively (**Figure 6c**; **Supplementary Table 13**). Even for EUR cohorts for which the largest same-ancestry GWAS was available, trans-ancestry PRS outperformed EUR-PRS (one-sided paired Wilcoxon test *P* = 3.3 × 10^-4^; **Extended Data Figure 9**).

Finally, we compared the performance of trans-ancestry PRS in each population. The best performance was found in the EUR cohorts (mean *R^2^* = 0.059; **Figure 6c**; **Supplementary Table 13**). Strikingly, the PRS explained around half of the heritability by the non-MHC common variants, which is the theoretical upper limitation (**Supplementary Table 2**). Even without the MHC region, we were able to identify 9.9% of the EUR population with an inherited genetic predisposition that conferred three times increased risk for RA (**Figure 6d**; **Methods**). The performance in the EAS cohorts was comparable with the EUR cohorts; the mean *R^2^* was 0.057 (Wilcoxon test *P* = 0.67 compared with EUR cohorts), and we were able to identify 5.5% of the EAS population with three times increased risk (**Figure 6c** and **6d**). However, we observed poor PRS performances in AFR, SAS, and ARB cohorts; the mean *R^2^* was 0.018 (Wilcoxon test *P* = 0.002 compared with EUR cohorts), and we were able to identify only 0.62% of these populations with three times increased risk (**Figure 6c** and **6d; Extended Data Figure 10**). Together, trans-ancestry PRS exhibited the best performance in all ancestries in this study. However, the PRS performance in each ancestry was substantially affected by its sample size in this trans-ancestry GWAS, which firmly claims that it is imperative to increase the sample sizes of underrepresented ancestries to equalize genetic predictability of disease status.

## DISCUSSION

This study identified 34 novel genetic signals and less than ten 95% credible sets at 43 loci. By using multiple functional annotations and prior immunological knowledge, we provided their potential molecular consequences. In addition to the novel loci, our comprehensive analyses provided novel biological interpretations to the known loci (e.g., the *IL6R* and *PADI4* loci). We conducted detailed analyses on ancestry specificity; although most genetic signals are shared across ancestries, we observed some ancestry-specific signals. We also found several candidates of critical TFs contributing to RA biology. This trans-ancestry study thus substantially advanced our understanding of RA biology.

We utilized the molecular QTL database to infer risk allele’s gene regulatory function. This approach is a standard approach but has a limited ability to explore the allelic role comprehensively, as reported in a previous study^67^. Since gene regulation is highly cell-type or cell-state specific, we need to extend the QTL experimental conditions to overcome this limitation. Single-cell QTL analysis may also represent a promising strategy^68^. Another promising approach is inducing risk alleles in target cell populations using gene-editing techniques; previous studies reported the feasibility of this approach^69–71^. Future advance in functional genetics would improve the biological insight from our GWAS.

Our study had insufficient power to detect significant signals for seronegative RA outside of the MHC region due to a limited sample size (4,515 cases of seronegative RA). Although we observed shared genetic risks between them (**Extended Data Figure 6**), this analysis was restricted to the loci detected in all RA or seropositive RA. To unveil the specific etiology of seronegative RA further, we need cohorts that are larger and have better representation of seronegative RA.

Poor PRS performance in non-EUR ancestries is becoming one of the major challenges in human genetics. Conducting a trans-ancestry GWAS on a large scale is a promising strategy to mitigate this issue. Indeed, trans-ancestry PRS performance in EAS cohorts was comparable to those in EUR cohorts, demonstrating that this study mitigated inequality of genetic benefit at least partially (**Figure 6c**). However, this study was underpowered to detect specific signals in non-EUR and non-EAS ancestries, resulting in poor PRS performance in these ancestries. To overcome these limitations, we need further efforts to diversify GWAS and increase sample sizes of underrepresented ancestries as in other common complex diseases.

## Methods

### Study participants

We included 35,871 RA patients and 240,149 control individuals of EUR, EAS, AFR, SAS, and ARB ancestry from 37 cohorts (**Supplementary Table 1**). All RA cases fulfilled the 1987 American College of Rheumatology (ACR) criteria^72^ or the 2010 ACR/the European League Against Rheumatism criteria^73^, or were diagnosed with RA by a professional rheumatologist. Among 35,871 patients, seropositivity status was available for 31,963; 27,448 were seropositive and 4,515 were seronegative (**Supplementary Table 1**). We defined seropositivity as the presence of rheumatoid factor or anti-citrullinated peptide antibodies. When a seropositive and seronegative GWAS has less than 50 cases, we excluded it from this study since GWAS with too few samples produces unstable statistics. All cohorts obtained informed consent from all participants by following the protocols approved by their institutional ethical committees. We have complied with all relevant ethical regulations.

### Genotyping and imputation

Genotyping platform and all quality control (QC) parameters of each cohort were provided in **Supplementary Table 1**. For QC of samples, we excluded those with (i) low sample call rate, (ii) closely related individuals, and (iii) outliers in terms of ancestries identified by PCA using the genotyped samples and all 1KG Phase 3 samples. Since 1KG Phase 3 does not include ARB samples, we did not exclude individuals in ARB cohort based on ancestral outliers. For QC of genotypes, we excluded variants meeting any of the following criteria: (i) low call rate, (ii) low MAF, and (iii) low *P* value for Hardy Weinberg equilibrium (HWE). Post-QC genotype data of each cohort was pre-phased using Shapeit2 or Eagle2 software. For EUR, AFR, and ARB cohorts, we conducted imputation with Minimac3 or Minimac4 software using the 1KG Phase 3 reference panel. For EAS and SAS cohorts, we conducted imputation with Minimac3 or Minimac4 using a reference panel which were generated by merging 1KG Phase 3 panel and whole genome sequence (WGS) data of each population (**Supplementary Table 1**); we used WGS data of 1,037 Japanese^74^, 89 Korean^75^, 7 Chinese^76^ or 96 Malaysian individuals^77^. For QC of imputed genotype data, we excluded low imputation quality variants (imputation r^2^ < 0.3) from each cohort, excluded variants with minor allele count less than ten in the reference panel, and then we included variants which passed QC in at least five cohorts; we finally included 20,990,826 autosomal variants and 736,614 X chromosome variants. The genomic coordinate is according to GRCh37 in all analyses.

### PCA and UMAP using all GWAS participants

To assess the ancestral background diversity of all GWAS participants, we projected them into the same PC space based on their imputed genotype data. From variants which passed QC criteria in all 37 cohorts (imputation r^2^ ≥ 0.3), we first identified 12,196 independent imputed variants (r^2^ < 0.2 in EUR samples of 1KG Phase 3). Due to data access restrictions, we were not able to transfer raw imputed genotype data across different institutes, and hence we were not able to conduct one PCA using all individuals. Therefore, we first conducted PCA using these variants and all 1KG Phase3 samples, and calculated the loadings of each variant for top 20 PCs. We then calculated each individual’s PC scores using these loadings and imputed dosage of our GWAS samples. We further conducted UMAP by umap package in R using these top 20 PC scores (n_neighbors = 30 and min_dist = 0.8).

### Genome-wide association analysis

We conducted GWAS in each cohort by a logistic regression model using PLINK2 software. We included age, sex, and genotype PCs within each cohort as covariates (details of covariates were provided in **Supplementary Table 1**). We then conducted meta-analysis using all cohorts by the inverse-variance weighted fixed effect model as implemented in METAL (trans-ancestry GWAS). For ancestries with multiple cohorts, we similarly conducted meta-analysis within each ancestry (EUR-, EAS-, and AFR-GWAS). When the seropositivity status was available, we also conducted GWAS only using seropositive RA samples and controls. We defined a locus as a genomic region within ±1 Mb from the lead variant, and we considered a locus as novel when it did not include any variants previously reported for RA. For non-RA autoimmune diseases (systemic lupus erythematosus, systemic sclerosis, Sjögren’s syndrome, dermatomyositis, juvenile dermatomyositis, and polymyositis), we used ±0.5 Mb window from the lead variant. We defined reported variants as significant variants (*P* < 5 × 10^-8^) reported in the GWAS Catalog (https://www.ebi.ac.uk/gwas/; e104_r2021-10-06) and those reported in previous literatures which we searched manually. Since we need a unique analytical strategy for the MHC locus, we excluded the MHC region (chr6:25Mb-35Mb) from this study, which will be reported in an accompanying project.

We performed stepwise conditional analysis within ±1 Mb from the lead variant. We conducted the same logistic regression model but including the dosages of the lead variants (index variants in the first round of conditional analysis) as covariates in each cohort; when the lead variants did not exist in post-QC imputed genotype data of a cohort (imputation r^2^ ≥ 0.3), we exclude that cohort from the analysis. We then conducted meta-analysis using the same strategy, and identified the 2^nd^ lead variant. We repeated these processes by sequentially adding the identified lead variants as covariates until we did not detect any significant associations (*P* < 5 × 10^-8^).

### Estimation of heritability and bias in GWAS results

We estimated heritability and confounding bias in EUR- and EAS-GWAS results with S-LDSC (version 1.0.0) using the baselineLD model (version 2.1). For EUR-GWAS, we utilized LD scores calculated in EUR samples in 1KG Phase3. For EAS-GWAS, we utilized LD scores calculated in EAS samples in 1KG Phase3. Since LDSC required a large sample size in GWAS (typically > 5K individuals), we restricted this analysis to EUR- and EAS-GWAS. We estimated prevalence of RA was 0.5% in both ancestries.

### Fine-mapping analysis

We conducted fine-mapping analysis using approximate Bayesian factor (ABF) and constructed 95% credible set for each significant locus^35^. We used trans-ancestry GWAS results. We included all the 122 autosomal loci (*P* < 5.0 × 10^−8^). We calculated ABF of each variant according to equation (1):

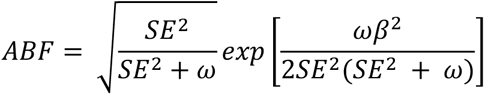

where *β* and *SE* are the variant’s effect size and standard error, respectively; *ω* denotes the prior variance in allelic effects (we empirically set this value to be 0.04)^78^. For each locus, we calculated posterior inclusion probability (PIP) of variant *k* according to equation (2):

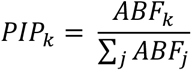

where *j* denotes all the variants included in the locus. We sorted all variants in order of decreasing PIP and constructed 95% credible set including variants from the top PIP until the cumulative PIP reached 0.95. When we compared the fine-mapping resolution across different GWAS setting, we also applied this fine-mapping strategy to EUR-GWAS and EAS-GWAS for all the 122 autosomal loci.

### Functional interpretation of fine-mapped variants

We quantified the enrichment of the 95% credible set variants at the 113 autosomal loci within ATAC-seq peaks in 18 hematopoietic populations using gchromVAR software^37^. We utilized the default parameters and ATAC-seq data processed by the developers. To access the specificity of a given ATAC-seq peak, we first normalized the read count of that peak in all 18 hematopoietic populations (each peak’s read count was divided by the total read counts, scaled by 1000,000, added 1 as an offset value, and log2-transformed), and transformed these 18 normalized counts into Z-scores.

### Functional interpretation of associated variants

We inferred the possible molecular consequences of all 148 variants detected in this study. We first focused on coding variants in LD with the lead variants in this GWAS (*r*^2^ > 0.6 both in EUR and EAS samples in 1KG Phase 3; when the lead variant is monomorphic in one ancestry, we only utilized the other ancestry). To annotate coding variants, we used ANNOVAR and assessed their potential impacts on protein function; we reported SIFT and Polyphen2 (HDIV) scores. To interpret their effects on gene regulation, we tested colocalization of our GWAS signals and eQTL or sQTL signals using coloc software^40^. We analyzed eQTL and sQTL results from Blueprint consortium database (CD4^+^ T cells, monocytes, and neutrophils) and eQTL results from v7 GTEx project database (48 tissues)^39,49^. Since coloc assumes GWAS and QTL signals are obtained from the same ancestry group, we only used EUR-GWAS results for this analysis. We defined GWAS and QTL signals are colocalizated when the posterior probability estimated by coloc software > 0.7.

### Capture RNA-seq of *PADI4* isoforms

We obtained total RNAs from THP-1 cells after stimulation with PMA for 72h, which induces the expression of *PADI4*^79^. We reverse-transcribed the RNA (10 ng) into cDNAs with Smart-seq2 primers^80^, and then amplified them by 10 cycles of polymerase chain reaction. We hybridized *PADI4* isoforms with xGen Lockdown Probes (5’-biotinylated 120-mer DNA probes synthesized by Integrated DNA Technologies) designed for all exons of *PADI4* main isoform. We captured the hybridized cDNAs with streptavidin-conjugated magnetic beads and then sequenced them with MinION sequencer using LSK-109 kit (Oxford Nanopore Technologies). We analyzed the sequenced reads with FLAIR (https://github.com/BrooksLabUCSC/flair).

We then performed sQTL analysis targeting *PADI4* using the eQTL data of peripheral blood subsets^5^. We reassembled and quantified the RNA-seq reads for *PADI4* isoforms including the newly discovered isoform using Cufflinks (http://cole-trapnell-lab.github.io/cufflinks/). We calculated the isoform ratio by dividing each isoform expression (FPKM) over total isoform expression. We used QTLtools (https://qtltools.github.io/qtltools/) for association testing.

### Stratified linkage disequilibrium score regression

We conducted stratified LD score regression (S-LDSC) to partition heritability. For this analysis, we used 707 cell-type-specific IMPACT annotations and 396 histone mark annotations^65,81^. IMPACT regulatory annotations were created by aggregating 5,345 epigenetic datasets to predict binding patterns of 142 transcription factors across 245 cell types. We computed annotation-specific LD scores using the EUR samples in 1KG Phase3 to analyze EUR-GWAS results. Similarly, we used EAS samples in 1KG Phase3 to analyze EAS-GWAS results. We estimated heritability enrichment of each annotation, while controlling for the 53 categories of the full baseline model. When we controlled the effect of an annotation, we conducted the same S-LDSC analysis but additionally including that annotation in a single model. We excluded variants in the MHC region (chr6:25 Mb-35 Mb).

### Trans-ancestry comparison of genetic signals

We first sought to compare the effect size estimates among GWAS results from each ancestry at the lead variants. However, the lead variants are not always the causal variants, and hence we restricted our targets to fine-mapped lead variants (*PIP* > 0.5). In addition, we excluded rare variants from this analysis because the effect sizes could not be accurately estimated for rare variants (MAF < 0.01 in either of the major ancestries in 1KG Phase 3). We finally included 30 fine-mapped variants for this analysis.

We next obtained trans-ancestry genetic-effect correlation using Popcorn software (version 1.0)^62^. We used summary statistics of EUR- and EAS-GWAS, and selected association statistics from variants with at least non-missing genotype from 5,000 individuals. We also excluded the MHC region from the analysis because of its complex LD structure. Using these post-QC summary statistics, we calculated the trans-ancestry genetic-effect correlation between EUR and EAS with precomputed cross-ancestry scores for EUR and EAS 1000 Genomes ancestries provided by the authors.

### Polygenic risk score

We used the pruning and thresholding method to calculate PRS in this study. We developed PRS models with six different conditions using combinations of two components; i) two variant selection settings used for PRS ((a) all variants or (b) variants within top 5% of the CD4^+^ T cell T-bet IMPACT annotation) and ii) three GWAS settings used for PRS (we used trans-ancestry, EUR-, or EAS-GWAS and refer PRS based on each of them as trans-ancestry, EUR-, or EAS-PRS, respectively). We designed our study so that the samples used in constructing PRS be independent from the samples in validation. When we evaluated the PRS performance in a given cohort, we re-conducted GWAS meta-analysis excluding that cohort to develop PRS models (**Figure 6a**). Before pruning, we removed rare variants from three GWAS results to reduce unstable effect estimates in PRS (MAF < 0.01 in EUR samples of 1KG Phase3 for EUR- and trans-ancestry GWAS; and MAF < 0.01 in EAS samples of 1KG Phase3 for EAS-GWAS). We also restricted this analysis to the variants which exist both in the GWAS results and post-QC imputed genotype of a cohort for which we apply PRS; and then we selected variants based on IMPACT annotation or utilized all variants (as described above). To LD-prune variants (*r^2^* < 0.2), we used haplotype information in EUR samples of 1KG Phase3 for EUR-and trans-ancestry GWAS and EAS samples of 1KG Phase3 for EAS-GWAS. For each of six conditions, we used 10 different *P* value thresholds: 0.1, 0.03, 0.01, 0.003, 0.001, 3.0 × 10^-4^, 1.0 × 10^-4^, 3.0 × 10^-5^, 1.0 × 10^-5^, and 5.0 × 10^-8^; we thus ended up having 60 different PRS models (6 conditions × 10 *P* value thresholds). We applied these 60 PRS models to 37 cohorts and applied a logistic regression model using per-individual PRS including the same covariates as used in GWAS; we evaluated PRS performances by Nagelkerke *R^2^*. In each of six PRS conditions, we selected the *P* value threshold with the largest average Nagelkerke *R^2^*, and used this *P* value threshold for the following analyses.

To discuss the PRS distribution in an ancestry, we first calculated PRS in each cohort using a specified condition; we next scaled those PRS values using the mean and the standard deviation of the PRS only of the control samples in that cohort; and we then merged PRS values across cohorts in an ancestry. We approximated the PRS distribution in general population by using that in control samples. For a given PRS value (at the right tail of PRS distribution), we compared the case-control ratios between individuals whose PRS is higher than that value and individuals whose PRS is lower than (or equal to) that value and calculated the odds ratio; and we then identified the minimum PRS value which showed odds ratio larger than three.

## Data Availability

The summary statistics and the PRS model with the best performance are publicly available at the following link:
https://data.cyverse.org/dav-anon/iplant/home/kazuyoshiishigaki/ra_gwas/ra_gwas-10-28-2021.tar.
The codes are available at our website:
https://github.com/immunogenomics/RA_GWAS.

https://data.cyverse.org/dav-anon/iplant/home/kazuyoshiishigaki/ra_gwas/ra_gwas-10-28-2021.tar

## Data availability

The summary statistics and the PRS model with the best performance are publicly available at the following link: https://data.cyverse.org/dav-anon/iplant/home/kazuyoshiishigaki/ra_gwas/ra_gwas-10-28-2021.tar.

The codes are available at our website: https://github.com/immunogenomics/RA_GWAS.

## Acknowledgement

We thank the Director of Health Malaysia for supporting the work described in the South Asian (SAS): the Malaysian Epidemiological Investigation of Rheumatoid Arthritis (MyEIRA) study. The MyEIRA study was funded by grant from Ministry of Health Malaysia (NMRR-08-820-1975) and the Swedish National Research Council (DNR-348-2009-6468). The GENRA study and the CARDERA genetics cohort genotyping were funded by Versus Arthritis (Grant Reference 19739 to ICS). The Nurses’ Health Study (NHS cohort) is funded by the NIH (R01 AR049880, UM1 CA186107, R01 CA49449, U01 CA176726, and R01 CA67262). The Swedish EIRA study was supported by the Swedish Research Council (to LK, LP and LA). SS was in part supported by The Mochida Memorial Foundation for Medical and Pharmaceutical Research, Kanae Foundation for the promotion of medical science, Astellas Foundation for Research on Metabolic Disorder, The JCR Grant for Promoting Basic Rheumatology, and Manabe Scholarship Grant for allergic and rheumatic diseases. ICS is funded by the NIHR Advanced Research Fellowship (Grant Reference NIHR300826). The views expressed are those of the author(s) and not necessarily those of the NIHR or the Department of Health and Social Care. KAS is supported by the Sherman Family Chair in Genomic Medicine and by a Canadian Institutes for Health Research Foundation grant (FDN 148457) and grants from the Ontario Research Fund (RE-09-090) and Canadian Foundation for Innovation(33374). S.Bae is supported by Basic Science Research Program through the NRF funded by the Ministry of Education (NRF-2021R1A6A1A03038899). RPK and JCE are funded by NIH (UL1 TR003096). CML is part-funded by the National Institute for Health Research (NIHR) Maudsley Biomedical Research Centre at South London and Maudsley NHS Foundation Trust and King’s College London. T.Arayssi was partially supported by the National Priorities Research Program (grant 4-344-3-105 from the Qatar National Research Fund, a member of Qatar Foundation). M.Kerick and JM are funded by Rheumatology Cooperative Research Thematic Network programme RD16/0012/0013 from the Instituto de Salud Carlos III (ISCIII, Spanish Ministry of Science and Innovation). YO is funded by JSPS KAKENHI (19H01021, 20K21834), AMED (JP21km0405211, JP21ek0109413, JP21ek0410075, JP21gm4010006, and JP21km0405217), JST Moonshot R&D (JPMJMS2021, JPMJMS2024), Takeda Science Foundation, Bioinformatics Initiative of Osaka University Graduate School of Medicine, Osaka University.

## Author contributions

K.Ishigaki, SS, C.Terao, YO, and S.Raychaudhuri conceived and designed the study. K.Ishigaki wrote the manuscript with critical inputs from SS, C.Terao, YL, YO, and S.Raychaudhuri. K.Ishigaki conducted meta-analysis and all GWAS downstream analyses with the help of SS, C.Terao, T.Amariuta, YL, YO, and S.Raychaudhuri. K.Yamaguchi and Y.Kochi conducted *PADI4* long-read sequencing and *PADI4* sQTL analysis. M.Koido, KT, Y.Kamatani, and C.Terao contributed to construction of population-specific reference panel. K.Ishigaki, SS, C.Terao, KS, VAL, ICS, SV, DP, JB, GX, JZ, CA, EK, RJC, KAS, M.Kerick, FM, M.Traylor, CML, HX, RS, T.Arayssi, JM, LK, YO, and S.Raychaudhuri conducted GWAS. C.Terao, CL.Too, VAL, SV, M.Takahashi, XW, LL, FL, DP, AB, GO, JB, SM, KPL, RJC, EWK, K.Matsuo, FM, SE, HX, K.Ikari, PKG, LP, YO, and S.Raychaudhuri contributed to genotyping experiments. C.Terao, KS, CL.Too, VAL, ICS, SV, KO, AM, MH, HI, M.Hammoudeh, SA, BKM, HH, HB, IWU, XW, LL, FL, DP, AB, GO, SMV, AJM, SH, HT, ET, AS, YM, Kenichi Yamamoto, SM, GX, JZ, CA, EK, GW, IvH, JC, KPL, RJC, HL, S.Bang, KAS, Nd, LA, S.Rantapää-Dahlqvist, EWK, S.Bae, RPK, JCE, XM, TH, PD, MS, M.Kerick, JCD, The Biobank Japan Project, K.Matsuda, K.Matsuo, TM, FM, KF, YT, AK, CML, SE, HX, RS, T.Arayssi, K.Ikari, M.Harigai, PKG, Kazuhiko Yamamoto, SLB, LP, JM, LK, YO, and S.Raychaudhuri contributed to collection of samples and management of genotype data and clinical information.

## Competing interests

The authors declare no competing interests.

## Notes

### Competing Interest Statement

The authors have declared no competing interest.

### Author Declarations

All cohorts obtained informed consent from all participants by following the protocols approved by their institutional ethical committees. We have complied with all relevant ethical regulations. IRB of participating institutions (including Brigham And Women's Hospital, Osaka University, and Riken) gave ethical approval for this work.

