## Supplementary note and figures for "Trans-ancestry genome-wide association study identifies novel genetic mechanisms in rheumatoid arthritis"

#### Comparison of fine-mapping resolution among different GWAS settings

To discuss the benefit of trans-ancestry study design in fine-mapping, we utilized three different GWAS settings; trans-ancestry (meta-analysis using all of five ancestries), EUR-, and EAS-GWAS. The 95% credible sets were smaller in the trans-ancestry GWAS than the EUR- and EAS-GWAS (one-sided paired Wilcoxon text $P = 1.1 \times 10^{-9}$ and $2.6 \times 10^{-18}$, respectively; **Supplementary Figure 3a**). Similarly, posterior inclusion probability (*PIP*) was higher in the trans-ancestry GWAS than the EUR- and EAS-GWAS (one-sided paired Wilcoxon text $P = 3.1 \times 10^{-11}$ and $6.1 \times 10^{-15}$, respectively; **Supplementary Figure 3b**).

To confirm the reason why trans-ancestry study design provided finer resolution results, we conducted a down-sampling experiment. We excluded 10 cohorts from EUR-GWAS and meta-analyzed them with similar-sized 10 cohorts from EAS, AFR, SAS, and ARB (down-sampled trans-ancestry GWAS; 22,253 cases and 73,080 controls in total); down-sampled trans-ancestry GWAS has almost identical sample size as in full sized EUR-GWAS (22,350 cases and 74,823). Even in this setting, the 95% credible sets were still smaller in the trans-ancestry setting (**Supplementary Figure 3c**). Similarly, *PIP* was still higher in the trans-ancestry setting (one-sided paired Wilcoxon text $P = 3.6 \times 10^{-5}$; **Supplementary Figure 3d**). These results suggested that the improved fine-mapping resolution in the trans-ancestry GWAS was not only due to the increase in sample size but also due to diversified LD structures.

#### Sample sizes required to detect ancestry-specific signals

In this study, we detected ancestry-specific signals only for EAS and EUR. Although ancestry-specific signals are relatively few, they include predominantly large effect size variants, many of which are missense, and hence they are valuable resources to understand the etiology of RA. Therefore, we sought to assess the sample size required to detect ancestry-specific signals. Since the true effect size and MAF of the causal variants are unknown, we assumed the lead variants of EUR-GWAS are the proxy of the causal variants. We selected 57 autosomal variants that passed genome-wide significance in EUR-GWAS, including three EUR-specific signals: rs2476601, rs34536443, and rs9826420 (see the definition in the main text). Using these variants, we conducted the power analysis and estimated the odds ratio corresponding to the sample size and MAF with the following conditions: power = 0.5, $\alpha$ = 5 x $10^{-8}$, and the case-control ratio = 0.23 (the actual ratio in EUR-GWAS) (**Supplementary Figure 4**). This analysis suggested that this study was underpowered to detect specific signals in non-EUR and non-EAS ancestries. For example, we need around 5,000 and 10,000 samples to detect equivalents of the *PTPN22* missense variant (rs2476601) and the *TYK2* missense variant (rs34536443) in non-EUR, respectively (**Supplementary Figure 4**).

**Effect size similarities between ancestries**

We examined the extent to which genetic effects are shared across ancestries. We focused on the 30 fine-mapped variants (*PIP* > 0.5; **Methods**). We compared effect sizes in EUR-GWAS with those in non-EUR-GWASs. We only found two variants with significant heterogeneities (**Supplementary Figure 5-7**); rs3093017 located at the *CCR6* locus (OR = 0.89 (0.87-0.92) in EUR-GWAS; OR = 0.82 (0.80-0.85) in EAS-GWAS; Cochran's Q test *P* value for heterogeneity ($P_{het}$) = 1.4 x $10^{-4}$ < 0.05 / (30 x 5)) and esv3585367 located at the *PADI4* locus (OR = 0.92 (0.90-0.95) in EUR-GWAS; OR = 0.83 (0.80-0.86) in EAS-GWAS; $P_{het}$ = 1.0 x $10^{-6}$ < 0.05 / (30 x 5)). Although rs3093017's effects were uniform within each ancestry, esv3585367 had a significant effect size heterogeneity within EAS cohorts ($P_{het}$ = 2.5 x $10^{-4}$); this remained significant even after restricting this analysis to seropositive RA ($P_{het}$ = 5.8 x $10^{-4}$) (**Supplementary Figure 6 and 7**; **Supplementary Table 4**). Therefore, these heterogeneous signals might suggest genetic differences among cohorts, rather than differences between EUR and EAS, which probably highlighting clinical heterogeneity in our GWAS. These results suggested that we need more efforts to collect clinically uniform samples to infer differences in genetic risk across ancestries.

**S-LDSC results using histone mark annotations**

In addition to IMPACT annotations, we analyzed 396 histone mark annotations and found significant enrichment in 55 annotations in either of EUR or EAS ($P < 0.05/396 = 1.3 \times 10^{-4}$; **Supplementary Table 11**); among them, the annotation of H3K4me1 in PMA-I stimulated primary $CD4^+$ T cells explained the largest fraction of heritability of EUR (52%; SE = 5.7%). However, controlling the effect of $CD4^+$ T cell T-bet IMPACT annotation cancelled out all significant enrichments of histone mark annotations (**Extended Data Figure 7**). Moreover, controlling the effect of H3K4me1 in PMA-I stimulated primary $CD4^+$ T cells was unable to cancel out significant enrichments of $CD4^+$ T cell T-bet annotation (**Extended Data Figure 7**). Therefore, we confirmed that $CD4^+$ T cell T-bet annotation can explain a large fraction of RA heritability and it outperforms histone mark annotations.

### Extended Data Figure

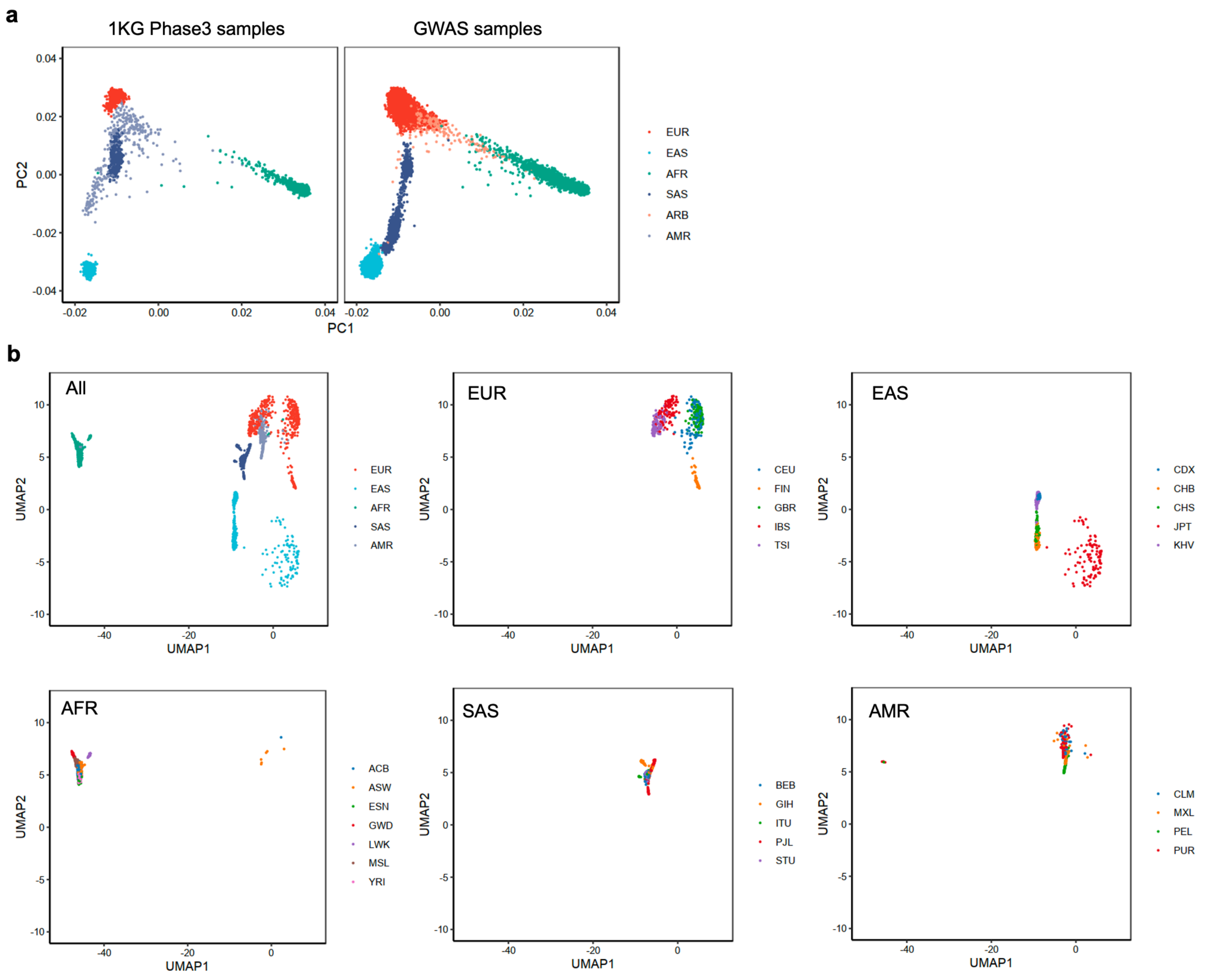


**Extended Data Figure 1. PCA and UMAP plot of 1KG Phase 3.**
**(a)** We projected each individual's imputed genotype into a PC space which was calculated using all individuals in 1KG Phase3.
**(b)** We further conducted UMAP analysis using the top 20 PC scores of all GWAS samples and all individuals in 1KG Phase3. We plotted all samples in 1KG Phase 3 or plotted samples in each ancestry separately. UMAP plot of all GWAS samples is provided in **Figure 1b**.

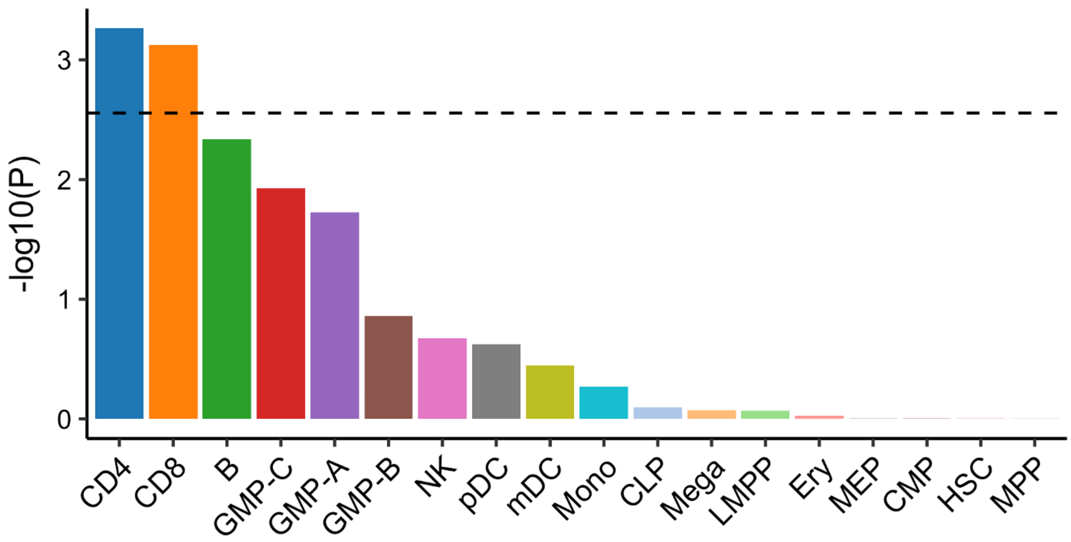


**Extended Figure Data 2. The enrichment of high *PIP* variants within open chromatin regions of 18 blood cell populations.**

The enrichment of high *PIP* variants within open chromatin regions of 18 blood cell populations was analyzed by gchromVAR software. The horizontal dashed line indicates Bonferroni corrected *P* value threshold (0.05/18=0.0027).

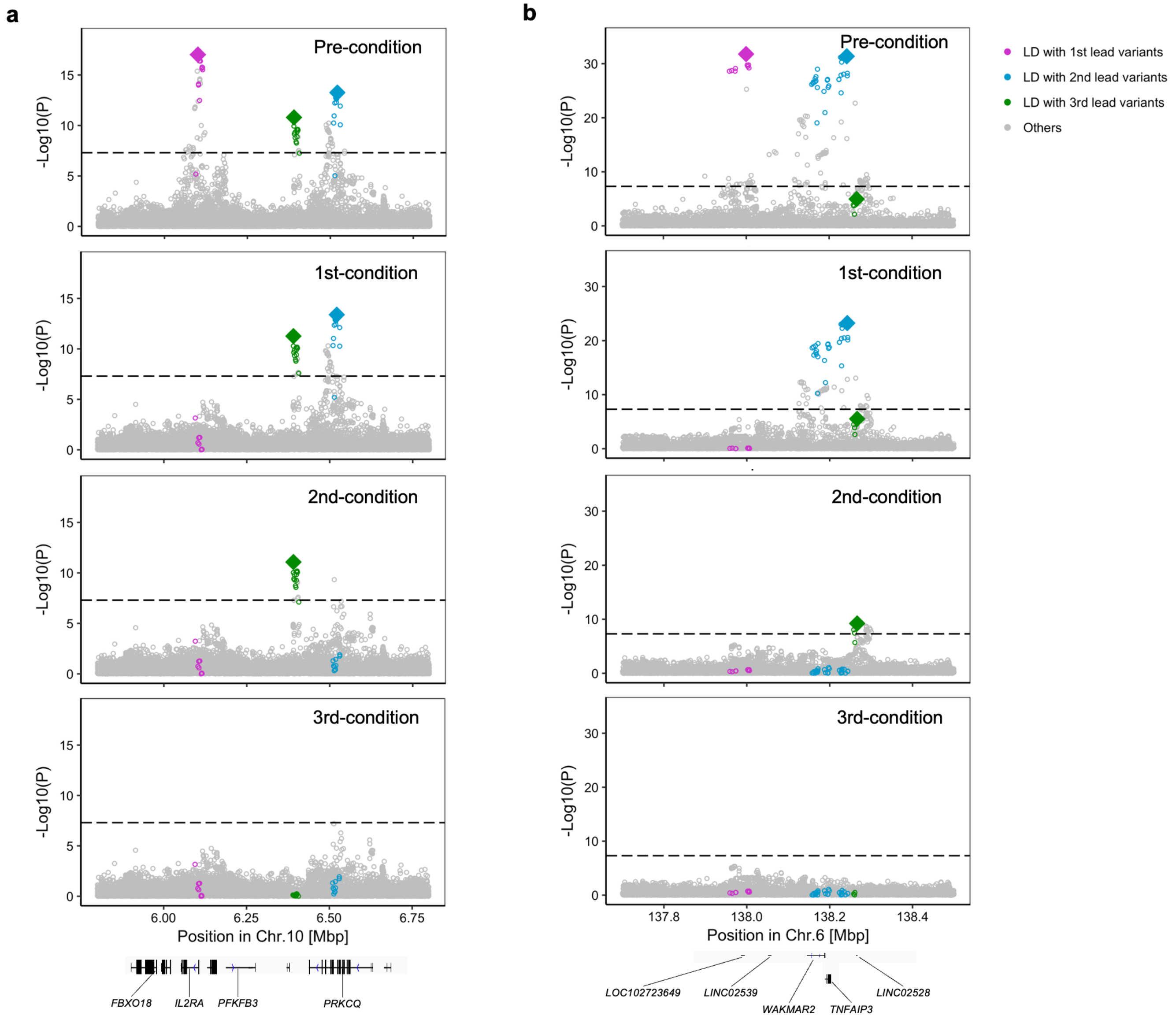


**Extended Data Figure 3. Conditional analysis results at the *IL2RA* and *TNFAIP3* loci.** Conditional analysis was conducted in each cohort and the results were meta-analyzed using the inverse-variance weighted fixed effect model (**a**, the *IL2RA* locus; **b**, the *TNFAIP3* locus). We used trans-ancestry GWAS results. The results at the *TYK2* locus were provided in **Extended Data Figure 5.** Variants in LD from the lead variant ($r^2$ > 0.6 both in EUR and EAS ancestries) in each round of conditional analysis are highlighted by different colors.

| Haplotype | rs12126142 | rs4341355 | Freq (case) | Freq (ctrl) |
|---|---|---|---|---|
| Hap 1 | Risk | Risk | 0.402 | 0.376 |
| Hap 2 | Risk | Protective | 0.215 | 0.224 |
| **Hap 3 (reference)** | **Protective** | **Risk** | **0.382** | **0.398** |
| Hap 4 | Protective | Protective | 0.0007 | 0.0019 |

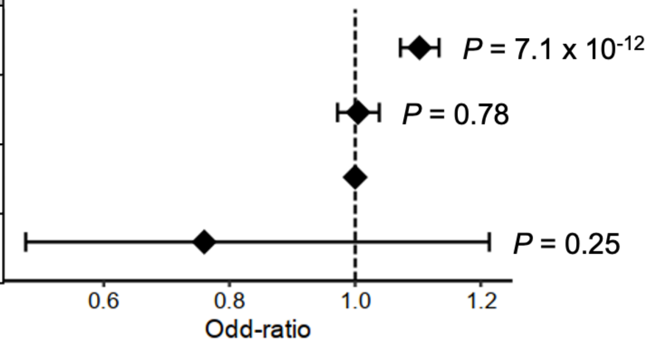


**Extended Data Figure 4. Haplotype-level association test at the *IL6R* locus.**
Haplotype-level association results at the *IL6R* locus. We defined haplotypes as shown in the table and we estimated the dosage of four haplotypes using phased imputed genotype data. We conducted multivariate logistic regression analysis in each of EUR cohorts and the results were meta-analyzed.

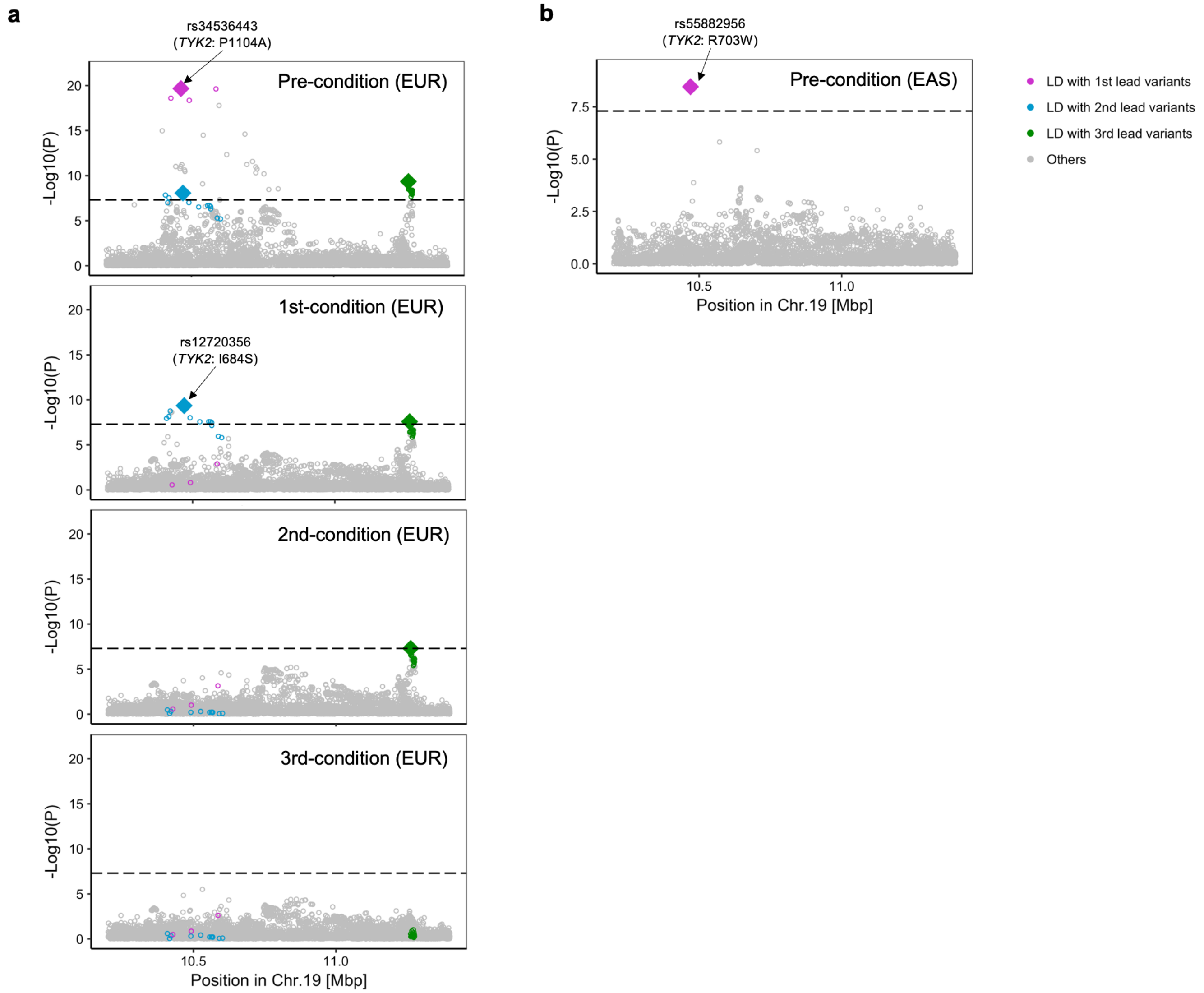


**Extended Data Figure 5. Three ancestry-specific signals at the *TYK2* locus.**
Conditional analysis was conducted in each cohort and the results were meta-analyzed using the inverse-variance weighted fixed effect model (**a**, EUR-GWAS; **b**, EAS-GWAS). Variants in LD from the lead variant ($r^2$ > 0.6 in EUR or EAS ancestries) in each round of conditional analysis are highlighted by different colors.

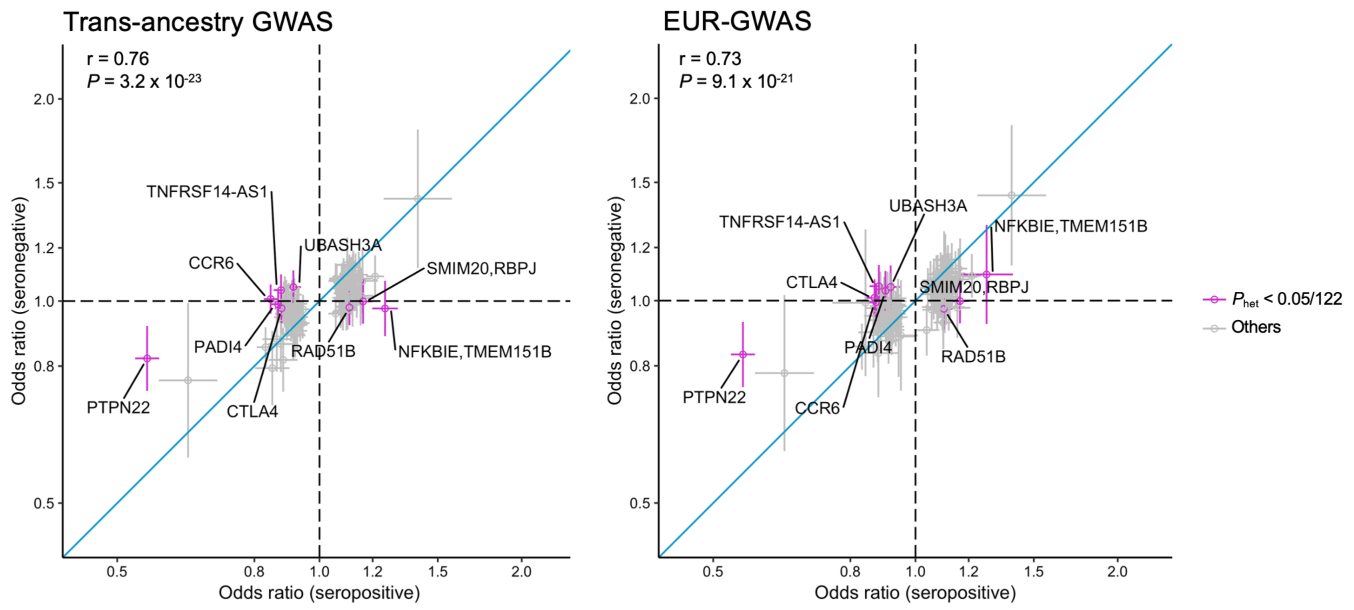


**Extended Data Figure 6. Effect size heterogeneity between seropositive and seronegative RA.** Odds ratio and its 95% confidence interval of seropositive and seronegative RA were plotted. Among the lead variants at 122 significant autosomal loci, we plotted 118 variants common in EUR of 1KG Phase 3 (MAF > 0.01). We provided the results from trans-ancestry GWAS results and EUR-GWAS. Effect size heterogeneity was assessed in the trans-ancestry GWAS results by Cochran's Q test ($P_{het}$). The Pearson's correlation data is provided.

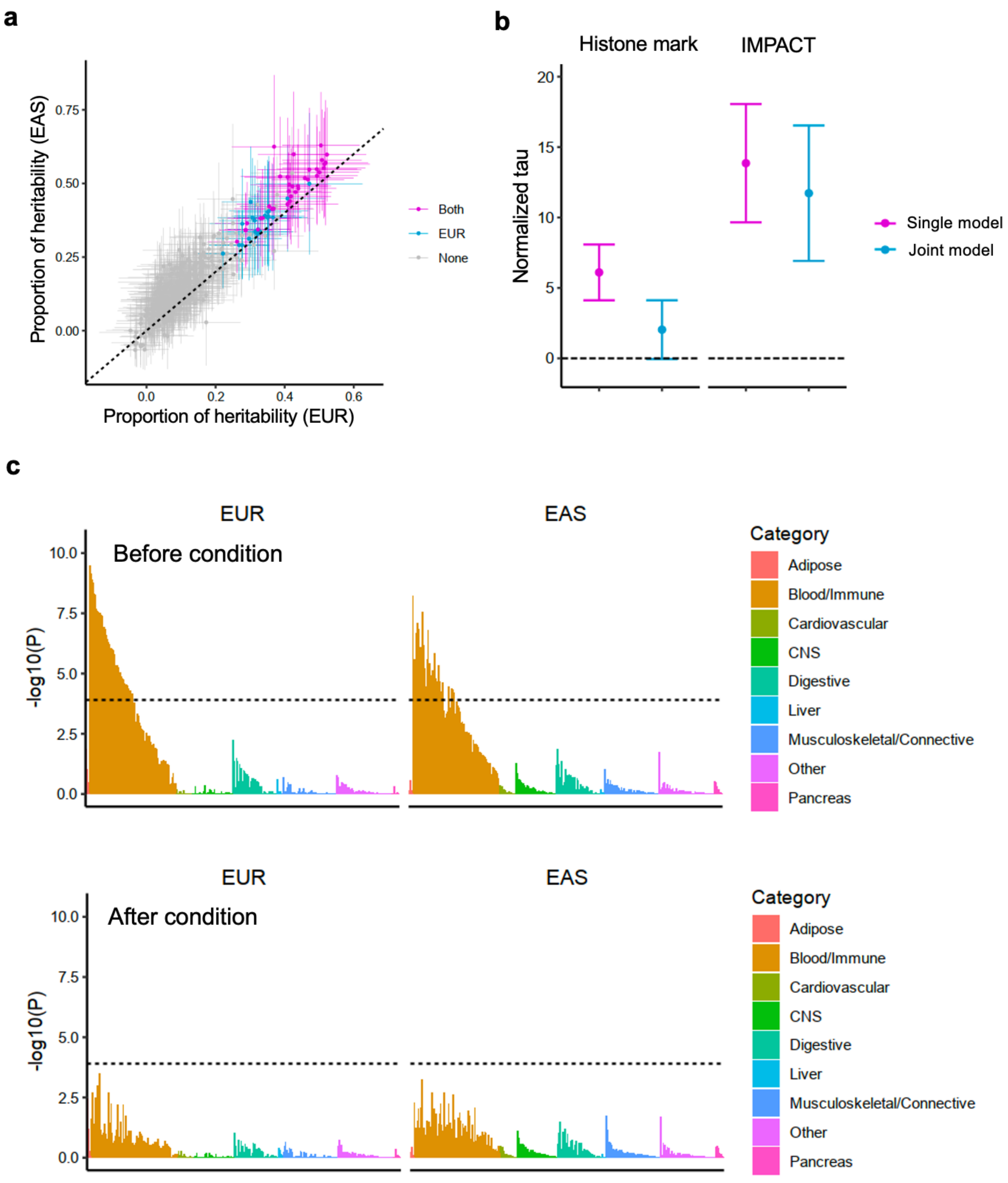


**Extended Data Figure 7. S-LDSC analysis using histone mark annotations.**
**(a)** The estimate and its 95% confidence interval of the heritability proportion explained by 396 histone mark annotations are provided. When a heritability enrichment is significant ($P < 0.05/396 = 1.3 \times 10^{-4}$), that annotation is colored by the type of GWAS.
**(b)** The best histone mark annotation (H3K4me1 in PMA-I stimulated primary $CD4^+$ T cells) and the best IMPACT annotation ($CD4^+$ T cell T-bet) were jointly modeled in S-LDSC analysis. Tau estimate (per variant heritability in each annotation) normalized by total per variant heritability and its 95% confidence interval are provided. EUR-GWAS results were used.
**(c)** *P* value indicates the significance of non-zero tau (per variant heritability) within each annotation. Each histone mark annotation is colored by its cell type category. Horizontal dashed line indicates Bonferroni-corrected *P* value threshold ($0.05/396 = 1.3 \times 10^{-4}$). Top panel shows the results without controlling the effect of the best IMPACT annotation ($CD4^+$ T cell T-bet) and the bottom panel shows the results with controlling this effect.

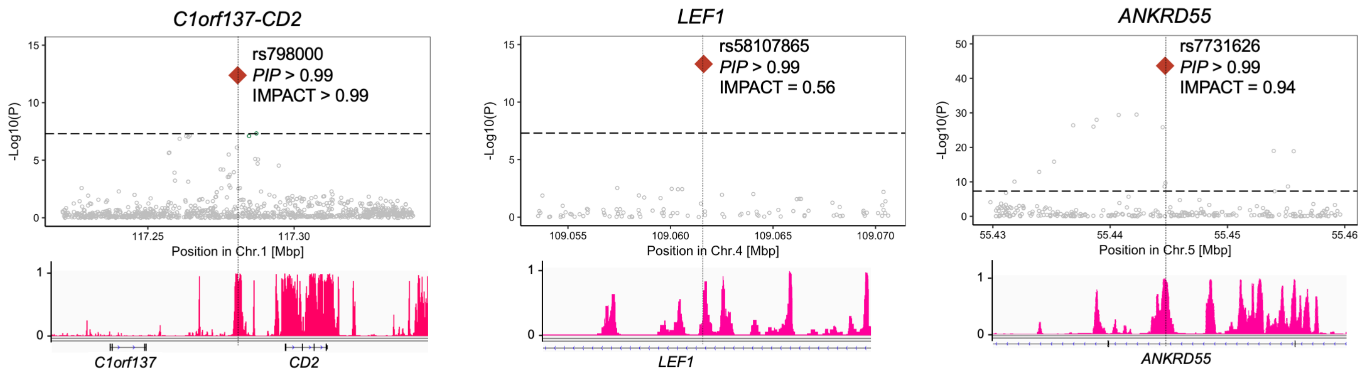


**Extended Data Figure 8. Fine-mapped variant with high IMPACT score.**
Among five loci whose lead variants exhibited high PIP and high IMPACT scores (both > 0.5), we showed three example loci with PIP > 0.99. *P* values in trans-ancestry GWAS are shown on the top panel. The track of CD4$^+$ T cell T-bet IMPACT annotation is shown on the bottom panel.

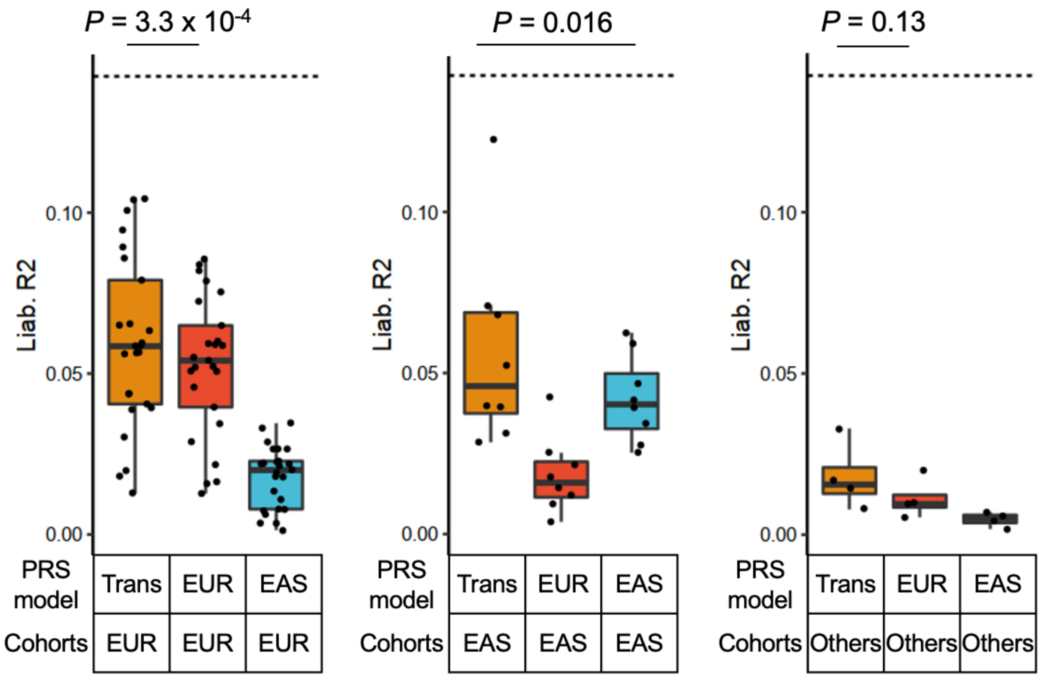


**Extended Data Figure 9. PRS performances in different ancestral groups.**
PRS performances (liability scale $R^2$) were provided for each combination of PRS models and cohort groups for which the PRS was applied. The differences between the group with the best performance and the second best were analyzed by Wilcoxon test. Within each boxplot, the horizontal lines reflect the median, the top and bottom of each box reflect the interquartile range (IQR), and the whiskers reflect the maximum and minimum values within each grouping no further than 1.5 x IQR from the hinge.

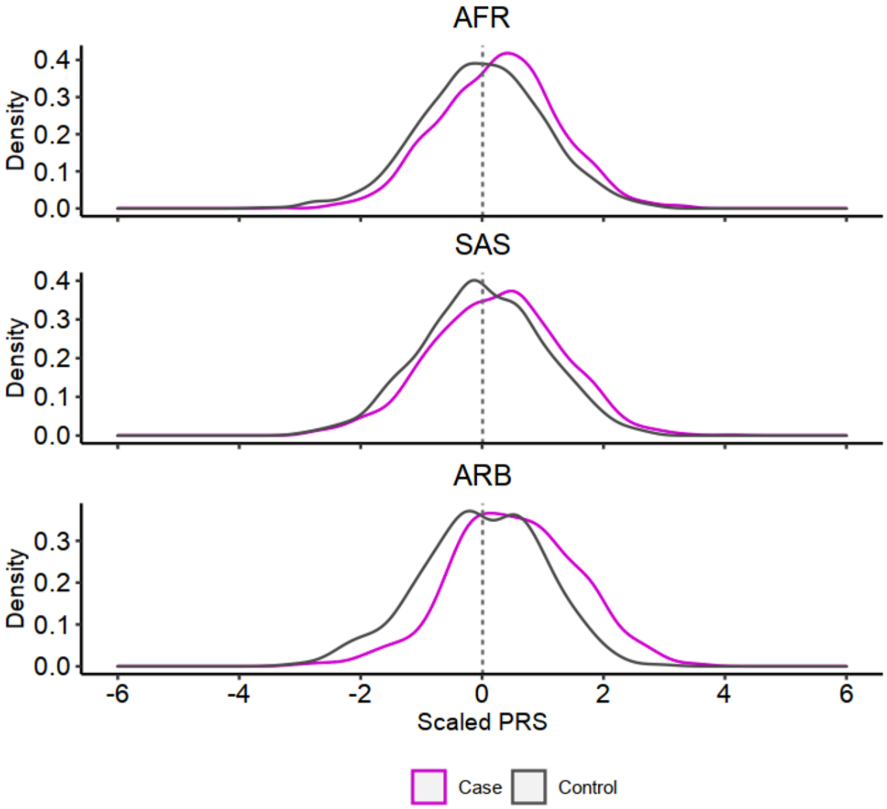


**Extended Data Figure 10. PRS distribution differences between case and controls.**
PRS distribution differences between case and controls. Trans-ancestry PRS with $CD4^+$ T cell T-bet IMPACT annotation was used. In each cohort, PRS was scaled using mean and SD of the control samples, and individual level data were merged across cohorts in an ancestry group.

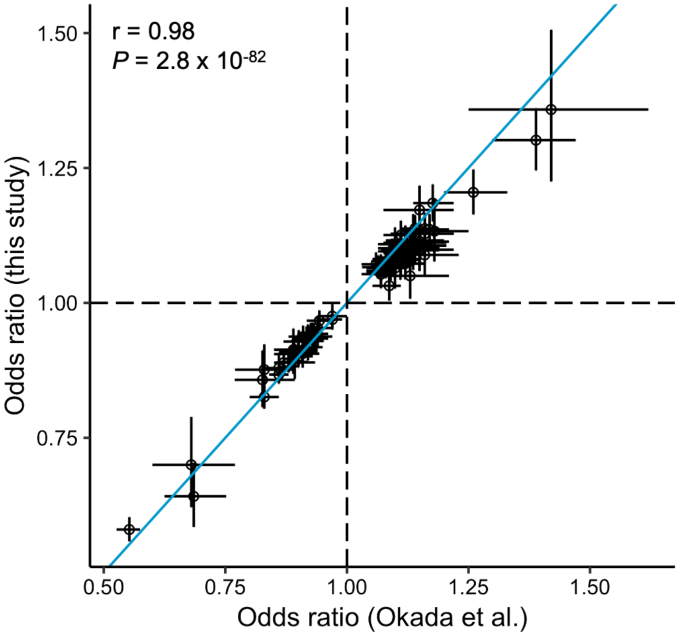


**Supplementary Figure 1. Comparison of effect sizes between this and previous GWAS.**
We compared effect size estimates between our study and a previous RA GWAS study at the 100 reported variants[1]. 6:149834574:T:C was excluded due to low imputation quality in our study. Odds ratios and their 95% confidence intervals are provided.

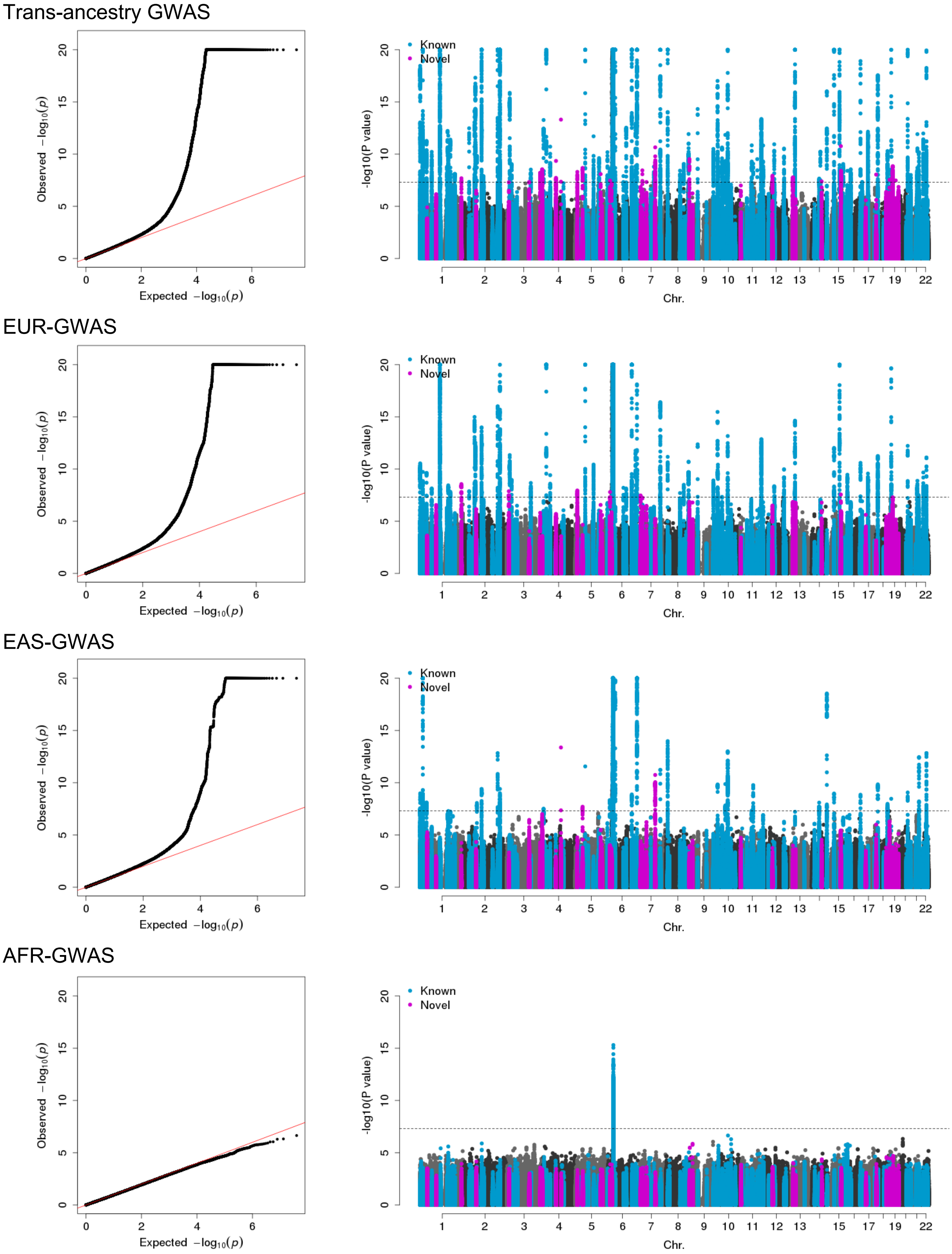
Trans-ancestry GWAS
Observed $-\log_{10}(p)$
Expected $-\log_{10}(p)$
-log10(P value)
Known
Novel
Chr.
EUR-GWAS
EAS-GWAS
AFR-GWAS

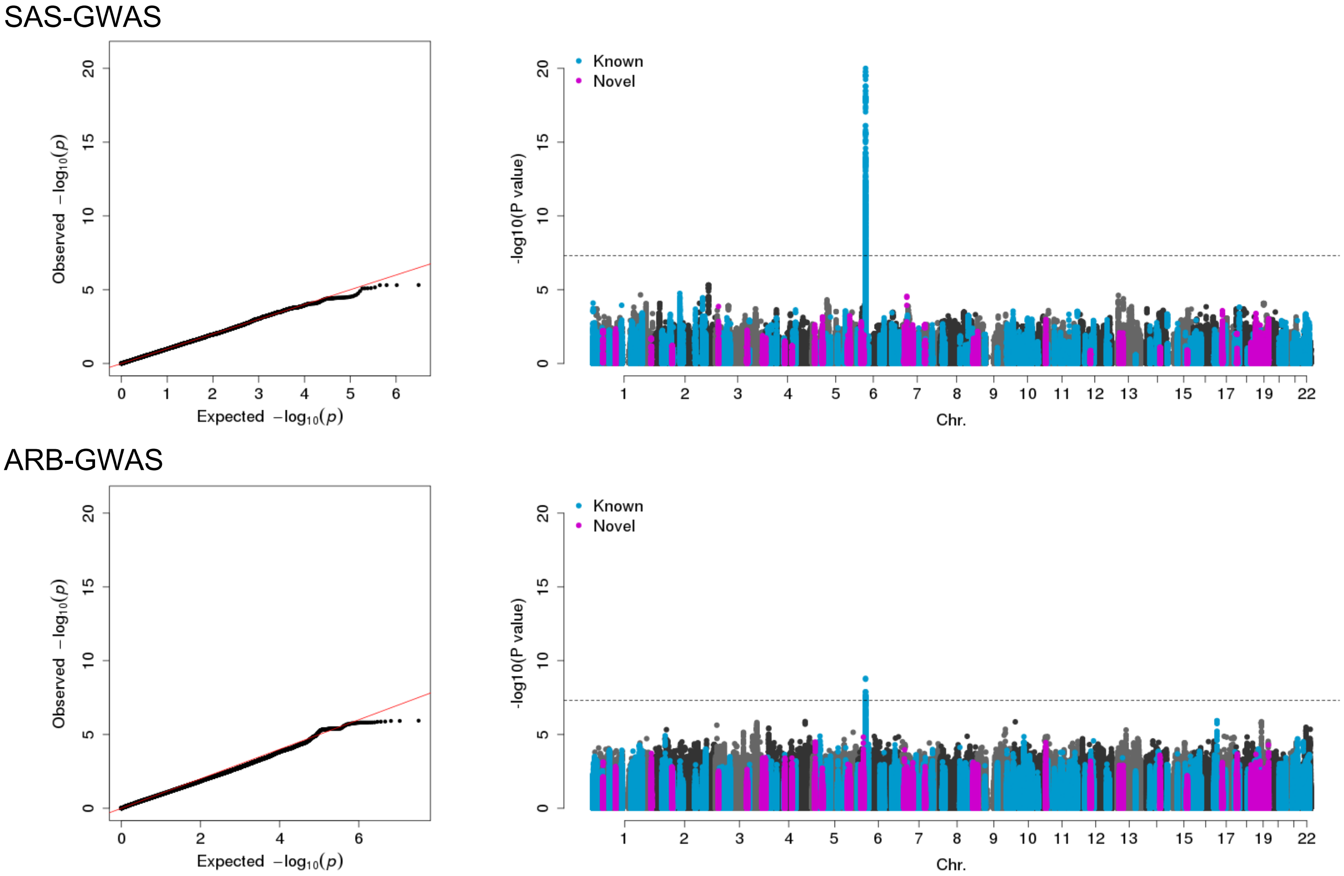


**Supplementary Figure 2. QQ plots and Manhattan plots for trans- and single-ancestry GWAS.**

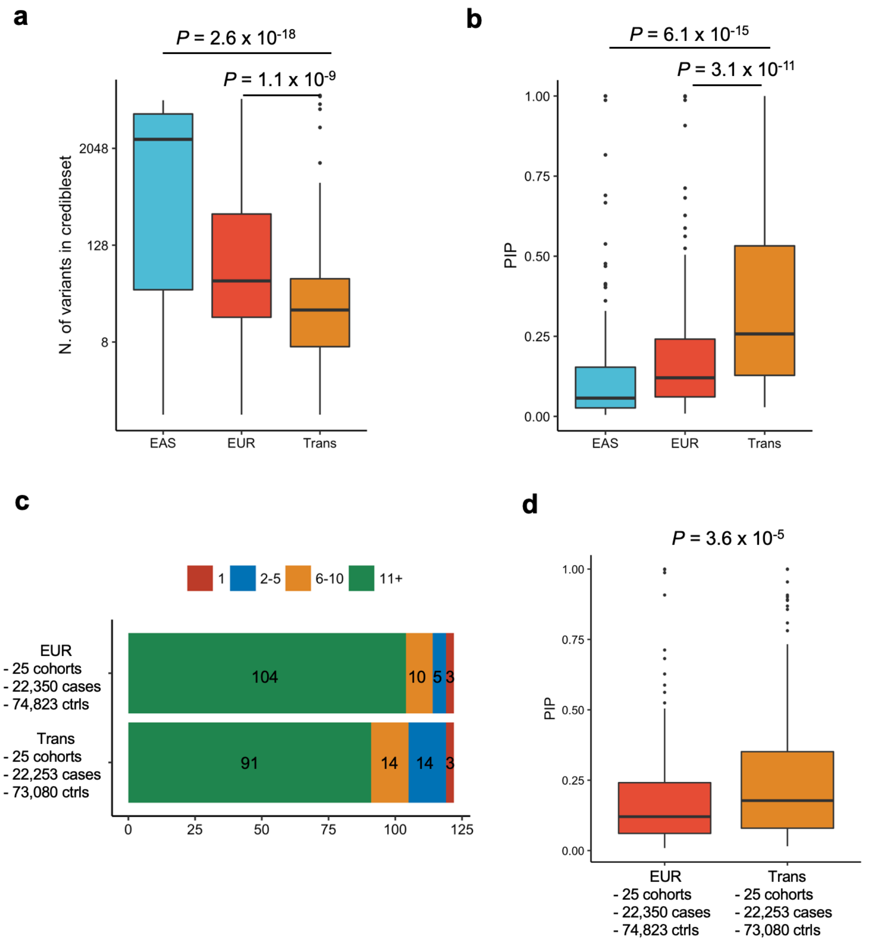


**Supplementary Figure 3. GWAS setting influences on fine-mapping resolution.**
**(a)** Number of variants in 95% credible set in three different GWAS settings at the 122 loci analyzed. The differences were assessed by one-sided paired Wilcoxon text.
**(b)** The *PIP* of the lead variant in three different GWAS settings at the 122 loci analyzed. The differences were assessed by one-sided paired Wilcoxon text.
**(c)** Among 122 loci analyzed, we counted the number of loci whose 95% credible set size was in a specified range. The results from full-sized EUR- and down-sampled trans-ancestry GWAS are provided.
**(d)** The *PIP* of the lead variant in two different GWAS settings at the 122 loci analyzed. The results from full-sized EUR- and down-sampled trans-ancestry GWAS are provided.
Within each boxplot, the horizontal lines reflect the median, the top and bottom of each box reflect the interquartile range (IQR), and the whiskers reflect the maximum and minimum values within each grouping no further than 1.5 x IQR from the hinge.

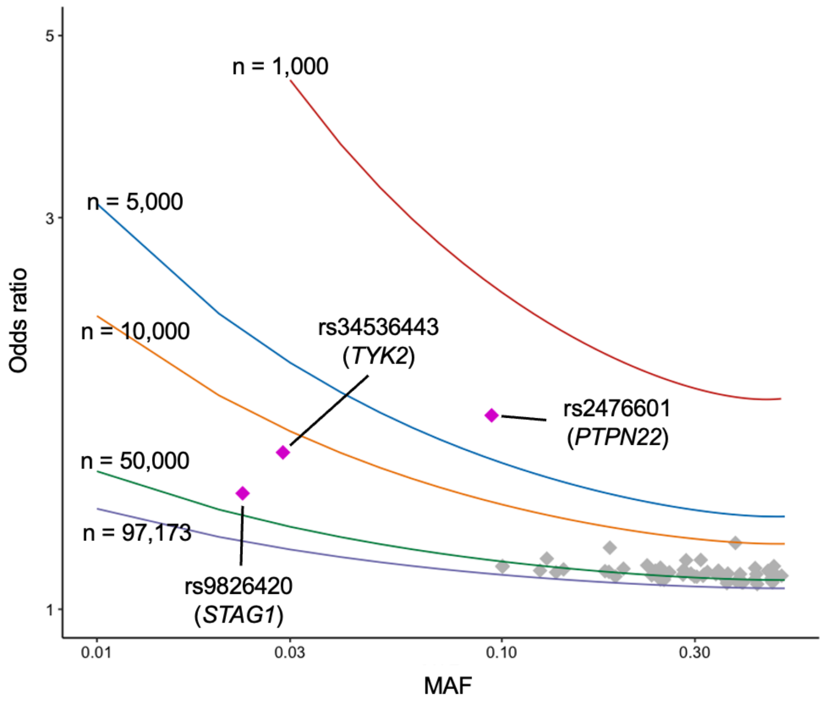


**Supplementary Figure 4. Sample sizes and the power of variant discovery.**
We selected 57 autosomal variants which passed genome-wide significance in EUR-GWAS, which include three EUR-specific signals: rs2476601, rs34536443, and rs9826420 (see the definition in the main text). Their odds ratios in EUR-GWAS and MAF in EUR are provided. The lines are the results of the power analysis that indicate the odds ratio corresponding to the sample size and MAF with the following conditions: power = 0.5, $\alpha = 5 \times 10^{-8}$, and the case-control ratio = 0.23 (the actual ratio in EUR-GWAS). The sample size of 97,173 is the actual size of this EUR-GWAS.

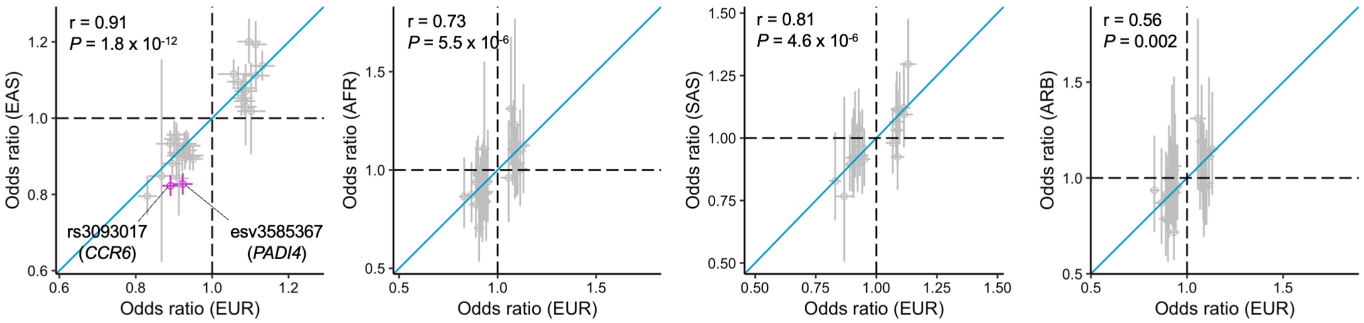


**Supplementary Figure 5. Shared GWAS signals across five ancestries.**
Odds ratio and its 95% confidence interval of EUR-, EAS-, AFR-, SAS-, and ARB-GWAS at 30 fine mapped variants (*PIP* > 0.5 and MAF greater than 1% in all of EUR, EAS, AFR, and SAS (1KG Phase 3)). rs3093017 and esv3585367 which had a significant heterogeneity in effect size estimate are highlighted by red (Cochran's Q test *P* value < 0.05 / (30 x 5)).

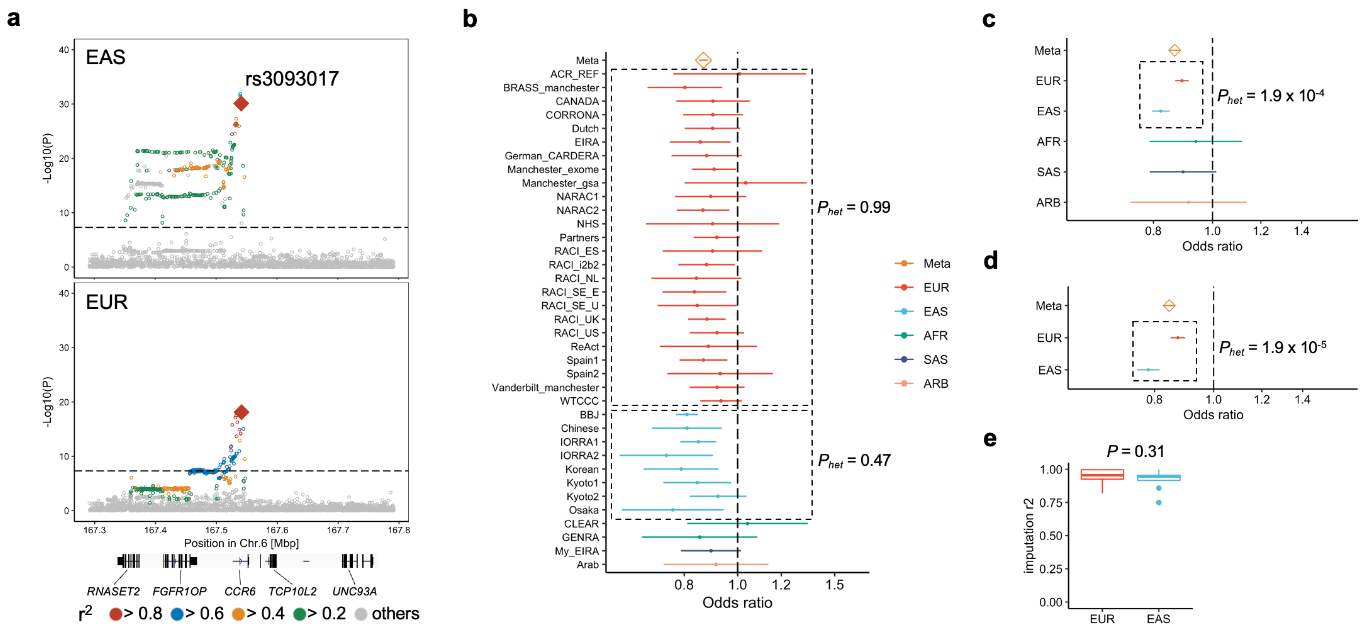


**Supplementary Figure 6. Heterogeneous GWAS signals at the *CCR6* locus.**

**(a)** GWAS *P* value plots at the *CCR6* locus. We used EAS- and EUR-GWAS results.

**(b)** Odds ratio and its 95% confidence interval of each cohort at the fine-mapped variant of the *CCR6* locus (rs3093017; *PIP* = 0.52). Effect size heterogeneity was analyzed by Cochran's Q test ($P_{het}$).

**(c)** Odds ratio and its 95% confidence interval of each ancestry at the fine-mapped variant of the *CCR6* locus (rs3093017; *PIP* = 0.52).

**(d)** same as (c) but cases were restricted to seropositive patients.

**(e)** Imputation quality ($r^2$) in EUR and EAS cohorts. The differences were assessed by Wilcoxon test.

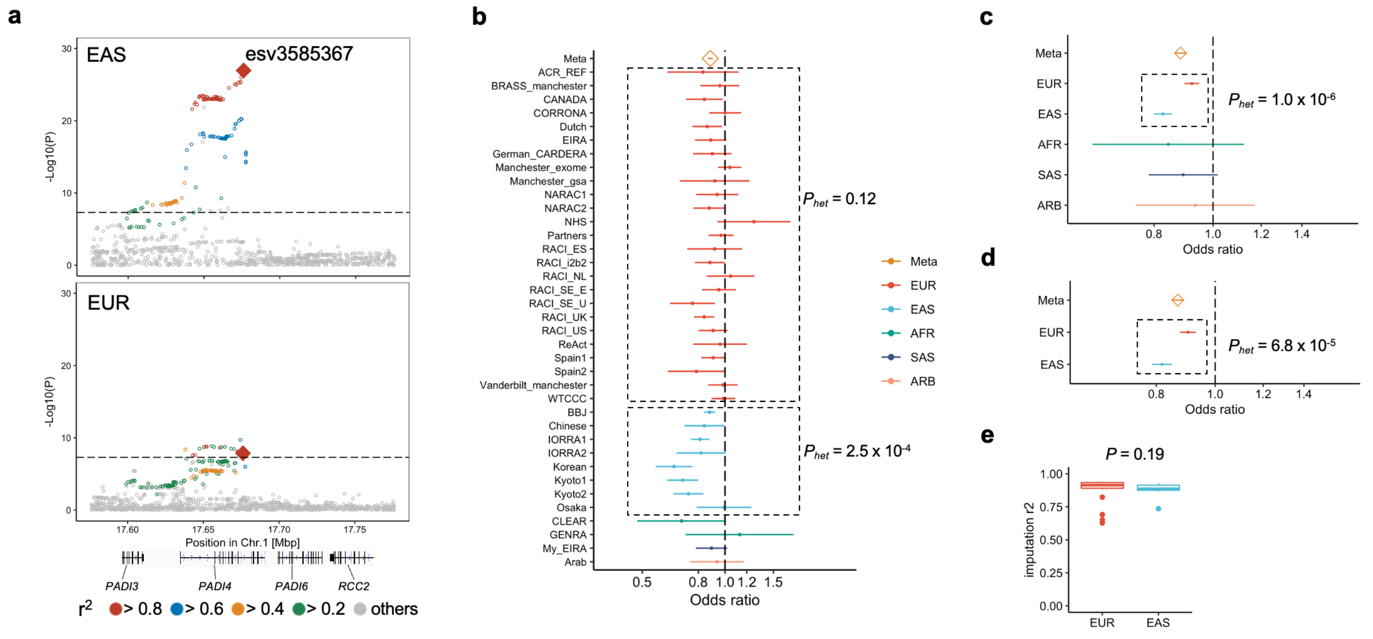


**Supplementary Figure 7. Heterogeneous GWAS signals at the *PADI4* locus.**
**(a)** GWAS *P* value plots at the *PADI4* locus. We used EAS- and EUR-GWAS results.
**(b)** Odds ratio and its 95% confidence interval of each cohort at the fine-mapped variant of the *PADI4* locus (esv3585367; *PIP* = 0.62). Effect size heterogeneity was analyzed by Cochran's Q test ($P_{het}$).
**(c)** Odds ratio and its 95% confidence interval of each ancestry at the fine-mapped variant of the *PADI4* locus (esv3585367; *PIP* = 0.62).
**(d)** same as **(c)** but cases were restricted to seropositive patients.
**(e)** Imputation quality ($r^2$) in EUR and EAS cohorts. The differences were assessed by Wilcoxon test.
